# Exploration of alcohol use disorder-associated brain miRNA-mRNA regulatory networks

**DOI:** 10.1101/2021.06.27.21259397

**Authors:** Yolpanhchana Lim, Jennifer E. Beane-Ebel, Yoshiaki Tanaka, Boting Ning, Christopher R. Husted, David C. Henderson, Yangfei Xiang, In-Hyun Park, Lindsay A. Farrer, Huiping Zhang

## Abstract

Alcohol use disorder (AUD) is due to gene expression changes in specific brain regions, but the underlying mechanism is not fully understood. We investigated AUD-associated miRNA-mRNA regulatory networks in multiple brain regions by analyzing transcriptomic changes in two sets of postmortem brain tissue samples and ethanol-exposed human embryonic stem cell (hESC)-derived cortical interneurons. miRNA and mRNA transcriptomes were profiled in 192 postmortem tissue samples (Set 1) from eight brain regions (amygdala, caudate nucleus, cerebellum, hippocampus, nucleus accumbens, prefrontal cortex, putamen, and ventral tegmental area) of 12 AUD and 12 control Caucasians. Nineteen differentially expressed miRNAs (fold-change>2.0 & *P*<0.05) and 97 differentially expressed mRNAs (fold-change>2.0 & *P*<0.001) were identified in one or multiple brain regions of AUD subjects. AUD-associated miRNA-mRNA regulatory networks in each brain region were constructed using differentially expressed and negatively correlated miRNA-mRNA pairs. AUD-relevant pathways (including *CREB Signaling, IL-8 Signaling*, and *Axonal Guidance Signaling*) were potentially regulated by AUD-associated brain miRNA-mRNA pairs. Moreover, miRNA and mRNA transcriptomes were mapped in additional 96 postmortem tissue samples (Set 2) from six of the above eight brain regions of eight AUD and eight control Caucasians, and some of the AUD-associated miRNA-mRNA regulatory networks were confirmed. Additionally, miRNA and mRNA transcriptomes were analyzed in hESC-derived cortical interneurons with and without ethanol exposure, and ethanol-influenced miRNA-mRNA regulatory networks were constructed. This study provided evidence that alcohol could induce concerted miRNA and mRNA expression changes in reward-related or alcohol-responsive brain regions. We concluded that altered brain miRNA-mRNA regulatory networks might contribute to AUD development.

## Introduction

Alcohol use disorder (AUD) is characterized by uncontrolled alcohol drinking due to physical and psychological dependence on alcohol. According to the 2019 National Survey on Drug Use and Health (NSDUH), AUD affects 14.1 million (4.2%) adult Americans (8.9 million men and 5.2 million women) [1]. Mounting evidence suggests that AUD is a complex genetic disorder, with an estimated heritability of about 50% [2]. Besides genetic variation, chronic alcohol consumption can lead to neuroadaptive phenomena, such as alcohol tolerance, dependence, and withdrawal [3]. The underlying molecular mechanisms of alcohol-induced neuroadaptations has not been fully explored, but it is believed that gene expression changes in specific brain regions are associated with AUD development.

Studies with animal-based models and human postmortem brains have demonstrated that alcohol exposure alters the expression of genes involved in diverse cellular functions. Using C57BL/6J mice as models, Ryabinin et al. observed expression changes of immediate early genes (c-fos, fosB, and zif268) in the hippocampus (HIP), the nucleus accumbens (NAc), the basolateral amygdala (AMY), and the lateral hypothalamus due to alcohol exposure [4]. Human postmortem brain studies have examined AUD-associated coding gene (or mRNA) expression changes in three brain regions [prefrontal cortex (PFC), NAc, and ventral tegmental area (VTA)] comprising the core reward circuitry. Differentially expressed coding genes identified in postmortem PFC of AUD subjects are potentially involved in transcription [5], aldehyde detoxification [6], nicotine response and opioid signaling [7], oxidative stress [5, 8], mitochondrial function [5, 6], myelination [9-12], calcium signaling [11], protein trafficking [10], fatty acid metabolism [6], cell cycling [13], cell adhesion [12], and neuronal apoptosis [5, 11, 12]. Differentially expressed coding genes identified in postmortem NAc of AUD subjects may participate in synaptic transmission [5, 8], vesicle formation and cell architecture [5], transcription and lipid metabolism [14], and oxidative phosphorylation, mitochondrial dysfunction and cytokine signaling [15]. Only one study is known to have examined mRNA transcriptomic changes in postmortem VTA of AUD subjects, and the identified differentially expressed coding genes likely contribute to neurotransmission and signal transduction [8]. These findings suggest that altered expression of coding genes or mRNAs in reward-related brain regions may underlie alcohol-induced neuroadaptations.

AUD-associated mRNA expression changes can only partially explain the molecular mechanisms of AUD. Noncoding RNAs, particularly small noncoding microRNAs (miRNAs), have drawn much attention as they are potent and multifunctional regulators of many biological processes. miRNAs are a class of about 22 nucleotide-long small noncoding RNAs that act as regulators of gene expression at the post-transcriptional level. They bind to the 3’ untranslated region (3’ UTR) of their target mRNAs, resulting in either mRNA degradation (when their sequences are perfectly matched) or translational inhibition (when their sequences are imperfectly matched) [16]. The function of miRNAs implies an additional layer of gene expression regulation besides genetic variation. Accumulating evidence suggests that alcohol could induce miRNA expression changes, leading to altered cellular functions. Expression changes in miRNAs and their target mRNAs have been demonstrated as a consequence of exposure of alcohol to cultured cells [17, 18] as well as mouse [19] and rat [20-23] brains. miRNA transcriptomic changes have also been observed in postmortem PFC [13, 24] and Nac [15] of AUD subjects by microarray-based transcriptome analysis.

Given that AUD is a genetically heterogeneous disorder, it is commonly agreed that multiple genes (including both coding and noncoding genes) and the interactions among them contribute to the etiology of AUD. Studies have shown that a single miRNA can target hundreds of mRNA transcripts while a single mRNA transcript can be simultaneously regulated by distinct miRNAs [25]. The particular role of miRNAs in posttranscriptional regulation implies that miRNAs fine-tune the expression of numerous genes involved in a variety of cellular functions and thus coordinate multiple cross-communicating pathways. To date, no studies are known to have explored AUD-associated miRNA-mRNA regulatory networks.

Here, we report the first network analysis of AUD-associated brain miRNAs and mRNAs. Specifically, we examined AUD-associated miRNA and mRNA transcriptomic changes in multiple brain regions of AUD subjects. We also performed miRNA-mRNA pairing analysis and constructed AUD-associated and brain region-specific miRNA-mRNA regulatory networks. To understand whether miRNA and mRNA expression changes in postmortem brains of AUD subject are due to alcohol consumption, we differentiated human embryonic stem cells (hESCs) into cortical interneurons and then used hESC-derived cortical interneurons as *in vitro* cellular models to examine ethanol-induced miRNA and mRNA transcriptomic changes. The convergence of multiple brain region transcriptome analysis and neuronal modeling could facilitate our understanding of the neuroadaptive mechanisms of AUD.

## Materials and methods

### Human postmortem brain tissues

Two sets of freshly-frozen autopsy brain tissue samples were obtained from the New South Wales Brain Tissue Resource Centre (NSWBTRC) in Australia. Set 1 included 480 [8 regions × (30 cases + 30 controls)] postmortem tissue samples dissected from eight brain regions [amygdala (AMY), caudate nucleus (CN), cerebellum (CRB), hippocampus (HIP), nucleus accumbens (NAc), prefrontal cortex (PFC), putamen (PUT), and ventral tegmental area (VTA)] of 30 (21 males and 9 females) AUD and 30 (21 males and 9 females) control subjects (NSWBTRC approved project #: PID409). Set 2 included 360 [6 regions × (30 cases + 30 controls)] postmortem tissue samples dissected from six brain regions (AMY, CN, CRB, HIP, PFC, and PUT) of 30 (20 males and 10 females) AUD and 30 (20 males and 10 females) control subjects (NSWBTRC approved project #: PID191). All subjects were Caucasian Australians with no history of illicit drug abuse or major psychotic disorders (such as schizophrenia and bipolar disorder) according to the criteria in the Diagnostic and Statistical Manual of Mental Disorder 4^th^ Edition (DSM-IV) [26]. Control subjects had no history of AUD.

### Isolation and selection of brain tissue RNA samples for miRNA and mRNA transcriptome analysis

Total RNAs were isolated from 10-50 mg of postmortem brain tissue samples using the miRNeasy Mini Kit (QIAGEN, Valencia, CA, USA). RNA integrity number (RIN) and concentration were measured using the Agilent 2100 Bioanalyser with the Agilent RNA 6000 Nano Kit (Agilent Technologies, Santa Clara, CA, USA). From the 480 Set 1 RNA samples, we selected 192 [from 8 brain regions of 12 AUD cases (6 males and 6 females) and 12 controls (6 males and 6 females)] with larger RINs (mean±SD: 6.6±1.3) for miRNA and mRNA transcriptome analysis. From the 360 Set 2 RNA samples, we selected 96 (from 6 brain regions of 8 male AUD cases and 8 male controls) with larger RINs (mean±SD: 5.9±1.4) for miRNA and mRNA transcriptome analysis. In both sets of selected RNA samples, cases and controls were matched by sex, age, RINs, and postmortem internals (PMIs). Characteristics (including the amount of daily alcohol use, sex, age, PMIs, RINs, brain weight, brain pH, cerebral hemispheres, smoking, and liver disease) of these two sets of RNA samples chosen for transcriptome analysis are summarized in **Supplementary Table S1**. Except the amount of daily alcohol consumption, other demographic variables were not significantly different in their measurements (or counts) between cases and controls.

### RNA-seq analysis of miRNA and mRNA transcriptomic changes in eight brain regions of AUD subjects (192 Set 1 RNA samples)

miRNA and mRNA expression profiles of the 192 selected Set 1 RNA samples were analyzed respectively by small RNA-seq and ribosome RNA (rRNA) depletion RNA-seq. Small RNA-seq was conducted as described in our previous study [27]. Briefly, small RNA libraries were generated using the NEBNext Multiplex Small RNA Library Prep Set for Illumina (Set 1) (NEB, Ipswich, MA, USA) with 250 ng of total RNAs. Purified cDNA libraries were pooled in equimolar ratios (12 libraries per pool) and multiplex sequenced at 1×75 bp on an Illumina HiSeq 2500 Sequencing System (Illumina, CA, USA). The Comprehensive Analysis Pipeline for miRNA Sequencing Data (CAP-miRseq) workflow [28] was used for raw reads (in fastq files) pre-processing, alignment, mature/precursor/novel miRNA qualification and prediction. The mean total number of reads per sample was 16,591,602, and the mean mapping rate (aligned reads/reads sent to Aligner) was 73.2%. Principal component analysis (PCA) of miRNA transcriptome data of these 192 samples (from 8 brain regions) showed clustered CRB and VTA samples, but samples from six other brain regions could not be separated by brain regions using the miRNA expression data (**Supplementary Fig. S1a**). The small RNA-seq fastq files and normalized read counts are available for downloading from the NCBI Gene Expression Omnibus (GEO) database (accession number: Pending).

Since most of the 192 selected postmortem brain RNA samples had RINs below 7 (**Supplementary Table S1**), the KAPA RNA HyperPrep Kit with RiboErase (KAPA Biosystems, Wilmington, MA, USA) was used to deplete ribosomal RNAs (rRNAs) and construct RNA-seq libraries with 1 μg of total RNAs as the starting material. Pooled libraries were loaded into individual lanes (80 pooled libraries/lane) of the NovaSeq S4 flow cell (Illumina, San Diego, CA, USA) by running the NovaSeq Xp workflow for 100 bp paired-end sequencing on a NovaSeq™ 6000 system (Illumina, San Diego, CA, USA). The bulk RNA-seq processing pipeline Pipeliner [29] was utilized to quantitate gene and isoform expression. The mean total number of reads per sample was 38,758,477, and the mean mapping rate (aligned reads/reads sent to Aligner) was 84.6%. PCA plotting of the mRNA-seq data showed similar sample clustering patterns as above using the miRNA-seq data (**Supplementary Fig. S1b**). The rRNA depletion RNA-seq fastq files and normalized read counts are available for downloading from the NCBI GEO database (accession number: Pending).

### Microarray analysis of miRNA and mRNA transcriptomic changes in six brain regions of AUD subjects (96 Set 2 RNA samples)

For miRNA transcriptome analysis, the Affymetrix GeneChip™ miRNA4.0 array (Affymetrix, Santa Clara, CA, USA) was used following the manufacturer’s instructions. This array was designed to detect all miRNAs in miRBase Release 20 [30]. It contains 30,424 probe sets for mature miRNAs of 203 species including 2,578 human mature miRNA probe sets, 2,025 human pre-miRNA probe sets, and 1,996 human snoRNA and scaRNA probe sets. Probe cell intensity files (or CEL files) for small noncoding RNAs (including miRNAs) were generated using the Affymetrix® GeneChip™ Command Console (AGCC) software. Small noncoding RNA CEL files were processed using the Affymetrix Expression Console (EC) software (v1.4.1) with the “MicroRNA Arrays – RMA (robust multi-array average) +DABG (detection above Background)-Human only” workflow as the default analysis for background adjustment and signal normalization as well as log_2_ transformation to create probe level summarization files (or CHP files). Quality control (QC) analysis of the CHP files was performed within the EC software, and the quality of the miRNA expression array data was visualized using box plots (**Supplementary Fig. S2a**). The CHP files for case and control samples were further analyzed by statistical programs to identify differentially expressed miRNAs and other small noncoding RNAs. The Affymetrix miRNA expression data has been deposited in the NCBI GEO database (accession number: Pending).

For mRNA transcriptome analysis, the Affymetrix Clariom™ D human array (Affymetrix, Santa Clara, CA, USA) was used following the manufacturer’s instructions. This array allows interrogating more than 540,000 transcripts (including coding and long non-coding genes, exons, and alternative splicing events as well as rare transcripts) using over 6.7 million probes. Probe cell intensity files (or CEL files) for transcripts were generated using the AGCC software. They were then analyzed using the Affymetrix EC software (v1.4.1) with the “Gene Level - RMA-Sketch (robust multi-array average with sketch quantile normalization)” workflow as the default analysis for background adjustment and signal normalization as well as log_2_ transformation to create probe level summarization files (or CHP files). Quality control (QC) analysis of the CHP files was performed within the EC software, and the quality of the mRNA expression array data was visualized using box plots (**Supplementary Fig. S2b**). The CHP files for case and control samples were further analyzed by statistical programs to identify differentially expressed mRNAs. The Affymetrix mRNA expression data has been deposited in the NCBI GEO database (accession number: Pending).

### Differentiation of hESCs into cortical interneurons and analysis of ethanol-induced miRNA and mRNA transcriptomic changes by RNA-seq

hESC-derived cortical interneurons were used as cellular models for analyzing ethanol-induced miRNA and mRNA transcriptomic changes. H1 hESCs (WiCell Research Institute, Madison, USA) were differentiated into cortical interneurons as previously described [31]. Briefly, H1 hESCs were dissociated with accutase (STEMCELL Technologies, Vancouver, Canada) and cultured in mTeSR1 (STEMCELL Technologies, Vancouver, Canada) on Matrigel (Corning Life Science, Tewksbury, USA) coated plate until 95% confluence. For neural induction (from Day 1 to Day 10), hESCs were cultured in the neural induction medium containing three inhibitors including 100 nM of LDN-193189 (Stemgent, Cambridge, MA, USA), 10 μM of SB-431542 (Tocris Bioscience, Bristol, United Kingdom), and 2 μM of XAV-939 (Stemgent, Cambridge, MA, USA), and the neural induction medium was changed daily. For ventral patterning (Day 11 - Day 18), the cells were cultured in neural induction media containing 100 ng/ml of SHH (R&D Systems, Minneapolis, MN, USA) and 1 μM of purmorphamine (Stemgent, Cambridge, MA, USA), and the medium was changed every other day. For final neuronal differentiation and maturation (Day 19 and after), the cells were cultured in the neuronal maturation medium supplemented with 20 ng/ml of BDNF (R&D Systems, Minneapolis, MN, USA), 200 μM of ascorbic acid (Sigma-Aldrich, St. Louis, MO, USA), and 200 μM of cAMP (Sigma-Aldrich, St. Louis, MO, USA), and the medium was changed every four days. After six weeks of maturation (totally 62 days *in vitro* differentiation), the H1 hESC-derived cortical interneurons were characterized by immunostaining (**Supplementary Fig. S3**) to confirm the expression of neuronal biomarkers as described in our previous study [32].

hESC-derived cortical interneurons were then cultured in the neuronal maturation media containing ethanol at a concentration of around 50-100 mM (equivalent to blood alcohol levels of heavy or intoxicated drinkers) for 7 days. The ethanol-containing neuronal maturation medium was changed every other day. After additional 24-hr culture without ethanol exposure, the cells were collected for total RNA isolation. Cell treatment experiments (exposed or unexposed to ethanol for 7 days) were performed in duplicate. Extra wells of cells treated with or without ethanol were fixed with 4% paraformaldehyde for cell morphology assay. Ethanol-exposed cells did not show apparent morphological changes (**Supplementary Fig. S4**).

miRNA transcriptomes of hESC-derived neurons (exposed or unexposed to ethanol) were profiled by small RNA-seq and the raw data obtained from small RNA-seq was processed by CAP-miRseq [28], as described above. The mean total number of reads per sample was 29,145,212, and the mean mapping rate (aligned reads/reads sent to Aligner) was 85.0%. The quality of the miRNA-seq data was visualized using box plots (**Supplementary Fig. S5a**). The miRNA-seq data has been deposited in the NCBI GEO database (accession number: Pending).

mRNA-seq was applied to profile the mRNA transcriptome of hESC-derived cortical interneurons since high-quality total RNA samples (RINs > 7) were extracted from cultured cells. The Illumina® TruSeq® Stranded mRNA Library Prep Kit (Illumina, San Diego, CA, USA) was used to construct mRNA-seq libraries. Pooled cDNA libraries (up to 8) were sequenced (2 × 100 bp) on the HiSeq 2000 system (Illumina, San Diego, CA, USA). The mRNA-seq raw data was processed by Pipeliner [29], as described above. The mean total number of reads per sample was 25,524,900 and the mean mapping rate (aligned reads/reads sent to Aligner) was 75.1%. %. The quality of the mRNA-seq data was visualized using box plots (**Supplementary Fig. S5b**). The mRNA-seq data has been deposited in the NCBI GEO database (accession number: Pending).

### Statistical analyses

Differential expression analysis was performed to identify differentially expressed miRNAs and mRNAs in each brain region of AUD subjects (given that gene expression is tissue-specific) and ethanol-exposed hESC-derived cortical interneurons. For RNA-seq count data from brain tissue samples, the differential expression analysis was performed using limma-voom [33], with a number of confounding factors being considered as covariates. We did principal component analysis (PCA) to extract the first three PCs for both technical (batch, RIN, and PMI) and biological (sex, age, brain weight, brain pH, left-right brain, smoking, and liver disease) confounding variables, and the obtained PC1, PC2, and PC3 were then used as covariates in the model matrix design for differential expression analysis.

### Bioinformatics analysis

The function of differentially expressed miRNAs was annotated using DIANA TOOLS – mirPath v.3 [34]. The gene ontology (GO) analysis of molecular functions (MF), biological processes (BP), and cellular components (CC) overrepresented in differentially expressed mRNAs was conducted using DAVID v6.8 [35]. Additionally, AUD-associated miRNA-mRNA pairs and their associated canonical pathways in each brain region were analyzed using the miRNA Target Filter function in Ingenuity Pathway Analysis (IPA, Ingenuity Systems, http://www.ingenuity.com). First, the differential expression analysis results [including folder changes (FC) and *P* values] of differentially expressed miRNAs identified in each brain region were uploaded as the input miRNA dataset for the IPA miRNA Target Filter function, and miRNA-mRNA pairs were then revealed using predicted miRNA-mRNA interactions from TargetScan [36], TarBase [37], and miRecords [38] as well as microRNA-related findings from peer-reviewed literature. Second, the differential expression analysis results (including FC and *P* values) of differentially expressed mRNAs identified in the same brain region were added, and the Expression Pairing function of the IPA miRNA Target Filter was applied to obtain miRNA-mRNA pairs in which their expression levels were negatively-correlated (i.e., upregulated miRNA-downregulated mRNA pairs or downregulated miRNA-upregulated mRNA pairs). Third, the obtained miRNA-mRNA pairs were used to construct miRNA-mRNA interaction networks. Finally, AUD-related canonical pathways were added to miRNA-mRNA networks to display miRNA-mRNA-pathway relationships.

## Results

### Differentially expressed miRNAs in multiple brain regions of AUD subjects and ethanol-exposed hESC-derived cortical interneurons

By small RNA-seq analysis of the 192 selected Set 1 samples (from 8 brain regions), we identified 19 differentially expressed mature miRNAs (absolute FC>2.0 & *P*<0.05) in one or more brain regions (4 in AMY, 5 in CN, 3 in CRB, 3 in HIP, 2 in NAc, 8 in PFC, 5 in PUT, and 2 in VTA) of AUD subjects (**Fig. 1** and **Supplementary Table S2**). Two miRNAs were upregulated (>2-fold increase & *P*<0.05) in multiple brain regions (miR-10a-5p: HIP and NAc; miR-144-3p: CN and PFC) of AUD subjects, while three other miRNAs were downregulated (>2-fold decrease & *P*<0.05) in multiple brain regions (miR-122-5p: AMY, CN, CRB, and VTA; miR-412-5p: AMY, CN, CRB, PUT, and VTA; and miR-6868-3p: AMY, CN, CRB, PFC, and PUT) of AUD subjects.

**Fig. 1.**
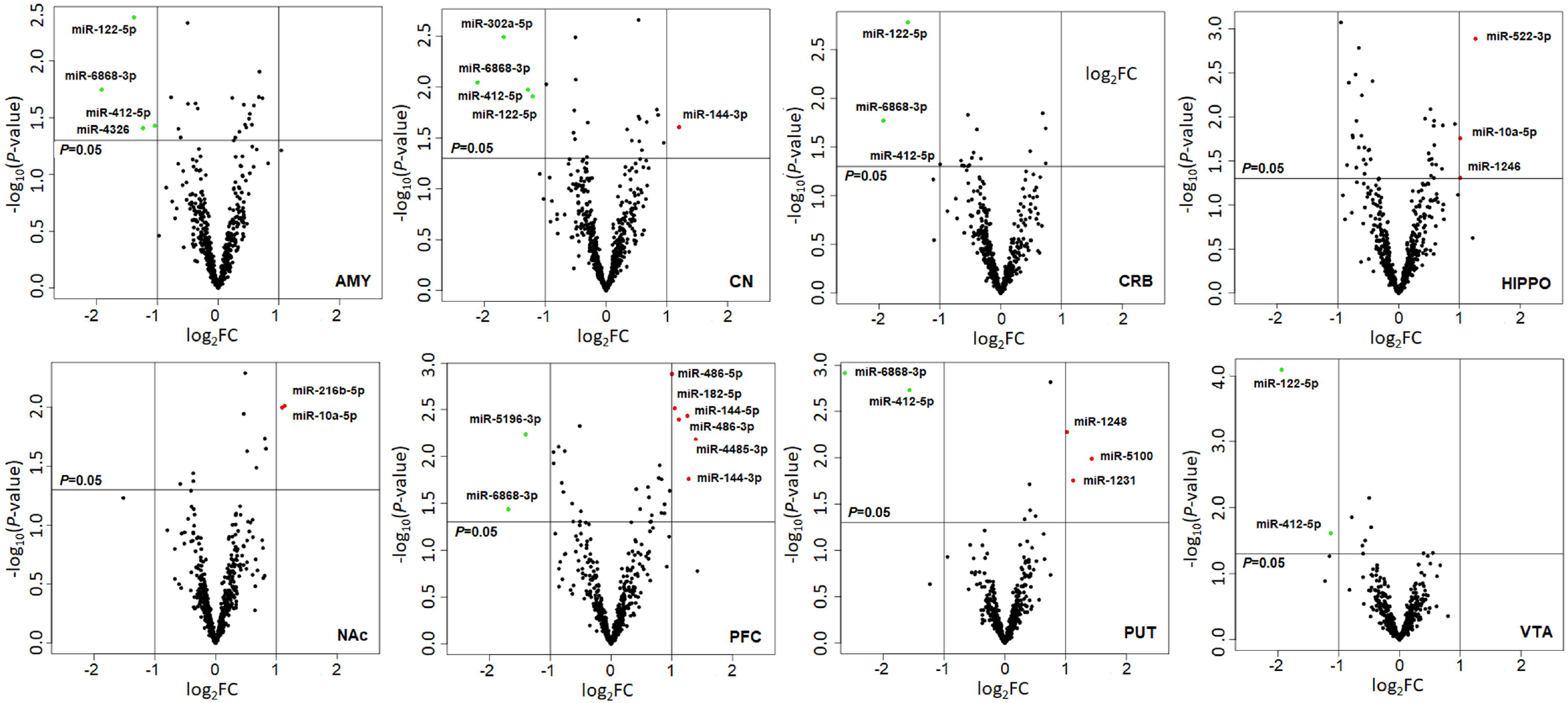
Volcano plots displaying differentially expressed miRNAs in eight regions of postmortem brains of subjects with alcohol use disorder (AUD) (the Set 1 sample). The vertical axis (y-axis) corresponds to the negative log_10_ of the *P*-value, and the horizontal axis (x-axis) displays the log_2_ of fold changes (FC). The red dots represent up-regulated miRNAs (log_2_FC > 1.0 & *P* < 0.05) and the green dots represent downregulated miRNAs (log_2_FC < -1.0 & *P* < 0.05). The horizontal line shows the *P*-value cutoff (*P* = 0.05) with points above the line having the *P*-value < 0.05 and points below the line having the *P*-value > 0.05. The two vertical lines indicate 2-fold changes. AMY: Amygdala; CN: Caudate Nucleus; CRB: Cerebellum; HIPPO: Hippocampus; NAc: Nucleus Accumbens; PFC: Prefrontal Cortex; PUT: Putamen; and VTA: Ventral Tegmental Area.

By Affymetrix miRNA 4.0 microarray analysis of the 96 selected Set 2 samples (from 6 brain regions), we identified 52 differentially expressed (absolute FC>2.0 & *P*<0.05) miRNAs in one or more brain regions (1 in AMY, 1 in CN, 3 in CRB, 4 in HIP, 44 in PFC, and 4 in PUT) of AUD subjects (**Supplementary Fig. S6 and Table S3**). Same as above, miR-412-5p was downregulated (>2-fold decrease & *P*<0.05) in five of the six brain regions (AMY, CN, CRB, HIP, and PUT) of AUD subjects.

We also examined ethanol-induced miRNA transcriptomic changes in hESC-derived cortical interneurons (as a cellular model) by small RNA-seq. A 7-day ethanol exposure led to differential expression of six miRNAs (absolute FC>2.0 & *P*<0.05) in hESC-derived cortical interneurons. Three miRNAs (miR-151b, miR-151a-5p, and miR-3135a) were upregulated (>2-fold increase & *P*<0.05), while three other miRNAs (miR-548bc, miR-3609, and miR-493-5p) were downregulated (> 2-fold decrease & *P* < 0.05) due to ethanol exposure (**Supplementary Fig. S7** and **Table S4**). The expression of miR-412-5p, which was downregulated in multiple brain regions of AUD subjects, was on a decreasing trend (1.3-fold decrease & *P*=0.263) in ethanol-exposed hESC-derived cortical interneurons.

Venn diagrams were made to display the number of differentially expressed miRNAs (*P*<0.05) shared between eight brain regions of AUD subjects (Set 1 and Set 2) and ethanol-exposed hESC-derived cortical interneurons (**Supplementary Fig. S8**). The *in vitro* cellular model study confirmed several AUD-associated brain miRNAs, including miR-98-3p (in AMY and HIP), miR-508-5p (in AMY), miR-548ah-3p (in CRB), miR-548p (in CRB), miR-486-5p (in PFC), miR-486-3p (in PFC), miR-139-3p (in PFC), and miR-151a-5p (in PFC).

### Differentially expressed mRNAs in multiple brain regions of AUD subjects and ethanol-exposed hESC-derived cortical interneurons

By rRNA depletion RNA-seq analysis of the 192 selected Set 1 samples (from 8 brain regions), we identified 97 differentially expressed (absolute FC>2.0 & *P*<0.001) coding genes (or mRNAs) in one or more brain regions (5 in AMY, 4 in CN, 21 in CRB, 11 in HIP, 4 in NAc, 46 in PFC, 11 in PUT, and 6 in VTA) of AUD subjects (**Fig. 2** and **Supplementary Table S5**). Three coding genes were upregulated (>2-fold increase & *P*<0.001) in multiple brain regions (*CHI3L1*: AMY, PUT, and VTA; *FSIP2*: CN, NAc, PFC, and PUT; and *MAFB*: CRB and VTA) of AUD subjects, while two other coding genes were downregulated (>2-fold decrease & *P*<0.001) in multiple brain regions (*CYYR1*: CN, CRB, and HIP; and *EDN3*: CN, HIP, NAc, and PUT) of AUD subjects.

**Fig. 2.**
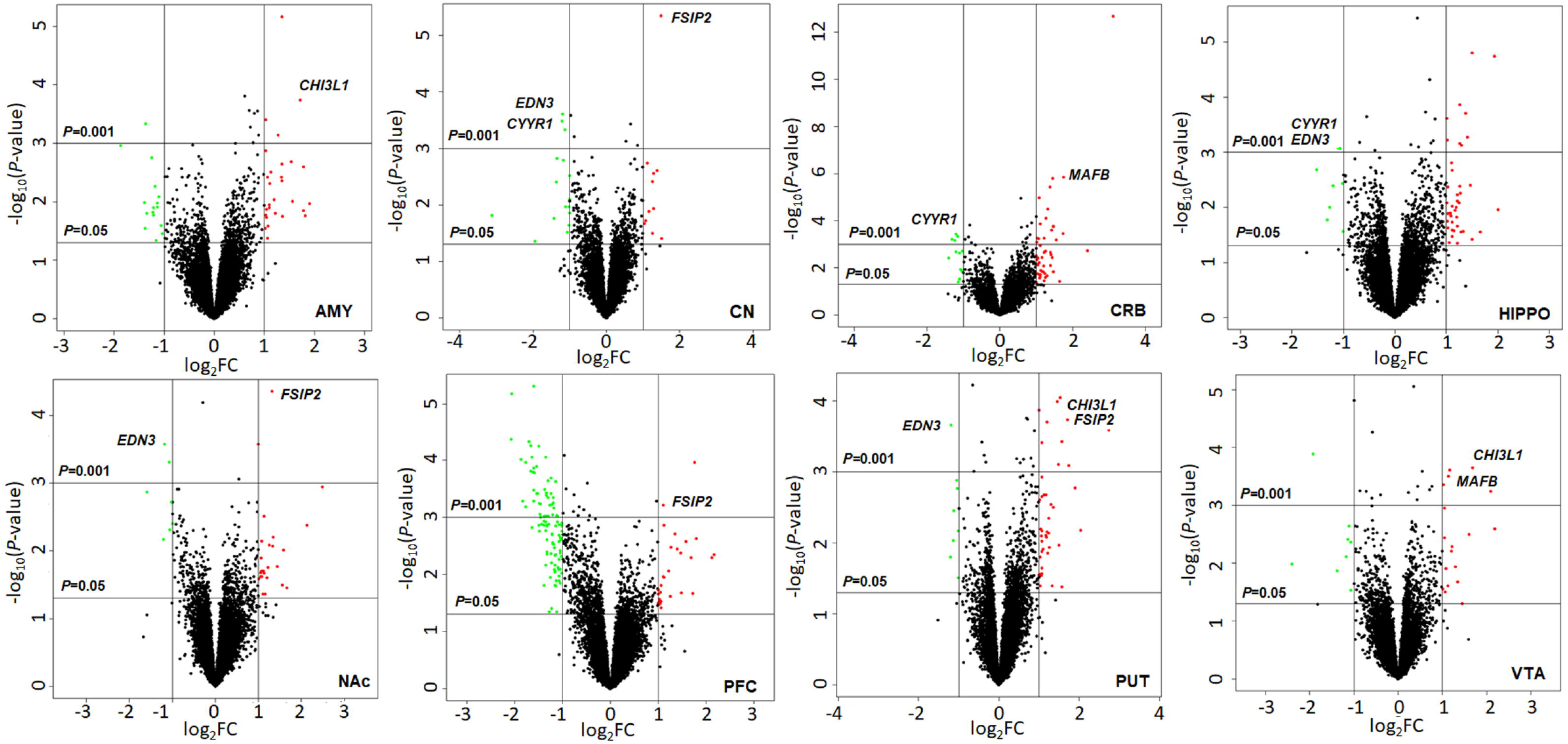
Volcano plots displaying differentially expressed mRNAs in eight regions of postmortem brains of subjects with alcohol use disorder (AUD) (the Set 1 sample). The vertical axis (y-axis) corresponds to the negative log_10_ of the *P*-value, and the horizontal axis (x-axis) displays the log_2_ of fold changes (FC). The red dots represent up-regulated mRNAs (log_2_FC > 1.0 & *P* < 0.05) and the green dots represent downregulated mRNAs (log_2_FC < -1.0 & *P* < 0.05). The horizontal line shows the *P*-value cutoff (*P* = 0.05 or 0.01) with points above the line having the *P*-value < 0.05 or 0.01 and points below the line having the *P*-value > 0.05 or 0.01. The two vertical lines indicate 2-fold changes. AMY: Amygdala; CN: Caudate Nucleus; CRB: Cerebellum; HIPPO: Hippocampus; NAc: Nucleus Accumbens; PFC: Prefrontal Cortex; PUT: Putamen; and VTA: Ventral Tegmental Area.

By Affymetrix Clarion D Human microarray analysis of the 96 selected Set 2 samples (from 6 brain regions), we identified six differentially expressed (absolute FC>2.0 & *P*<0.001) coding genes (or mRNAs) in one or more brain regions (upregulated *OCLN* and downregulated *GLS2* in AMY; downregulated *SLC47A1, PROX1, MYO5B*, and *TNC* in HIP; and upregulated *OCLN* in PUT) of AUD subjects (**Supplementary Fig. S9** and **Table S6**). Decreased expression of *TNC* was also observed in multiple brain regions of Set 1 AUD subjects (AMY: 2.53-fold decrease & *P* = 0.016; CN: 1.48-fold decrease & *P* = 0.307; HIP: 1.38-fold decrease & *P* = 0.406; and PFC: 2.61-fold decrease & *P* = 0.016).

We also examined ethanol-induced mRNA transcriptome changes in hESC-derived cortical interneurons by mRNA-seq. A 7-day ethanol exposure did not cause coding gene expression changes at the above significance level (absolute FC>2.0 & *P*<0.001) (**Supplementary Fig. S10**). When the significance level was set at FC>2.0 & *P*<0.01, 19 coding genes showed differential expression, and all of them were downregulated after a 7-day ethanol exposure (**Supplementary Table S7**).

Venn diagrams were used to show the number of differentially expressed mRNAs (*P*<0.05) shared between eight brain regions of AUD subjects (Set 1 and Set 2) and ethanol-exposed hESC-derived cortical interneurons (**Supplementary Fig. S11**). The *in vitro* cellular model study confirmed a number of AUD-associated mRNAs, including 15 mRNAs in the AMY, 15 mRNAs in the CN, five mRNAs in the CRB, five mRNAs in the HIP, seven mRNAs in the NAc, eight mRNAs in the PFC, 10 mRNAs in the PUT, and four mRNAs in the VTA. Among them, 13 AUD-associated coding genes identified in multiple brain regions were found differentially expressed in ethanol-exposed hESC-derived cortical interneurons (**Supplementary Fig. S11**).

### Functional annotations of miRNAs and mRNAs differentially expressed in the brain of AUD subjects

The function of the top 19 differentially expressed (absolute FC>2.0 & *P*<0.05) miRNAs (**Supplementary Table S2**) identified in one or more of the eight brain regions of AUD subjects (Set 1) was annotated by DIANA-mirPath. The top 14 KEGG pathways (*P*<0.0001), including *Morphine Addiction* (*P*=3.1×10^−8^; 15 miRNAs), *Cocaine Addiction* (*P*=1.5×10^−5^; 15 miRNAs), and *Amphetamine Addiction* (*P* = 1.0×10^−4^; 15 miRNAs), were associated with mRNAs potentially targeted by these 19 miRNAs (**Fig. 3**). The function of the top 97 differentially expressed (absolute FC>2.0 & *P*<0.001) mRNAs (**Supplementary Table S**5) identified in one or more of the eight brain regions of AUD subjects (Set 1) was annotated by DAVID. The gene ontology (GO) enrichment analysis showed that several molecular functions (MF; such as *Phosphatidate Phosphatase Activity*), biological processes (BP; such as *Central Nervous System Myelination*), and cellular components (CC; such as *Integral Component of Membrane*) were enriched in these 97 differentially expressed mRNAs. GO terms (MF, BP, and CC) over-represented (*P*<0.05) for this gene set are displayed in **Supplementary Fig. S12**.

**Fig. 3.**
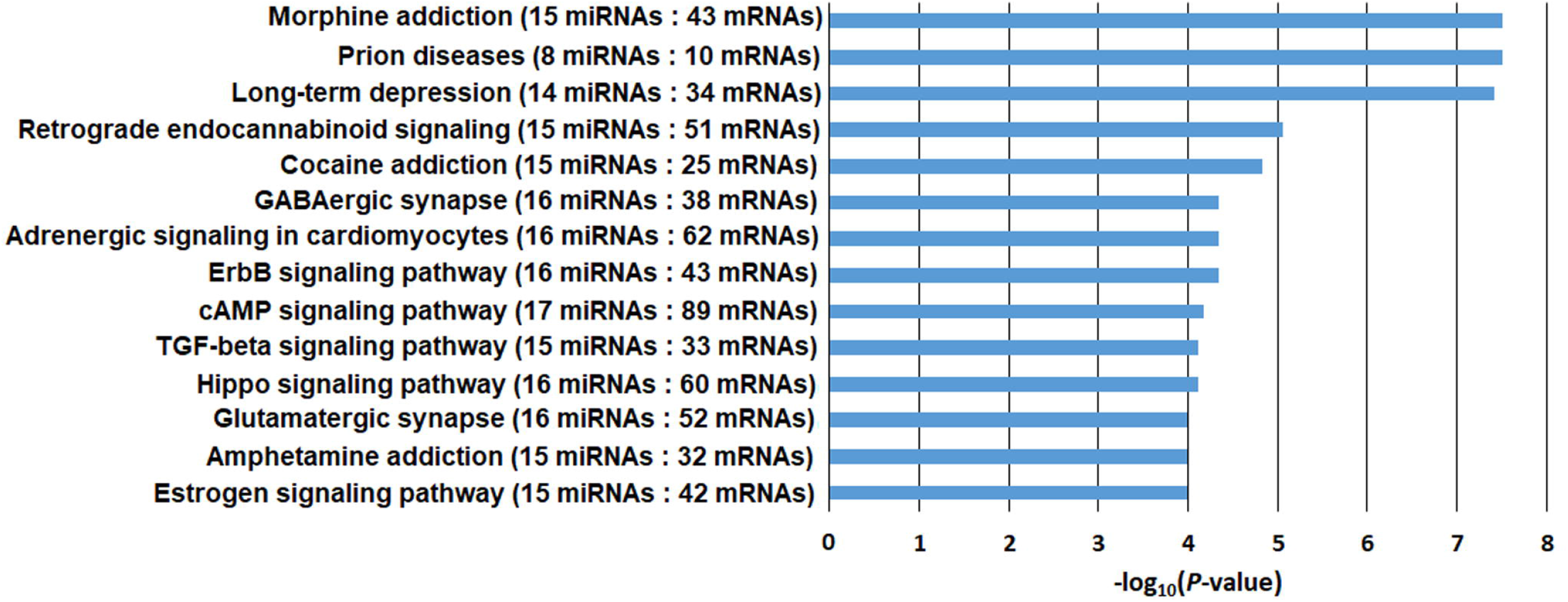
DIANA-mirPath KEGG pathway enrichment analysis of mRNAs potentially targeted by 19 differentially expressed miRNAs (absolute FC > 2.0 & *P* < 0.05) identified in one or multiple brain regions of subjects with alcohol use disorder (AUD) (the Set 1 sample). Numbers in parentheses: the number of differentially expressed miRNAs (absolute FC > 2.0 & *P* < 0.05) and the number of predicted target mRNAs involved in specific pathways. KEGG pathways with enrichment *P* values < 10^−4^ (or –log_10_*P* > 4.0) are listed.

### AUD-associated brain miRNA-mRNA regulatory networks

Differentially expressed and negatively correlated miRNA-mRNA pairs were included in brain region-specific IPA network analysis. The differential expression analysis *P* value was set at < 0.05 and the absolute FC was set at > 1.3 for both miRNAs and mRNAs. AUD-associated miRNA-mRNA-pathway networks for each of the eight brain regions (the Set 1 sample) are shown in **Figures 4** and **5**. Within the AMY, 13 miRNAs (6 upregulated and 7 downregulated) and 13 paired mRNAs (10 upregulated and 3 downregulated) could regulate four pathways (*CREB Signaling in Neurons, STAT3 Pathway, IL-8 Signaling*, and *Axonal Guidance Signaling*) (**Fig. 4a**). Within the CN, 10 miRNAs (6 upregulated and 4 downregulated) and eight paired mRNAs (3 upregulated and 5 downregulated) could regulate four pathways (*CREB Signaling in Neurons, Gap Junction Signaling, Axonal Guidance Signaling*, and *Neuroinflammatory Signaling*) (**Fig. 4b**). Within the CRB, seven miRNAs (2 upregulated and 5 downregulated) and nine paired mRNAs (8 upregulated and 1 downregulated) could regulate three pathways (*Synaptogenesis Signaling, CREB Signaling in Neurons*, and *Neuroinflammatory Signaling*) (**Fig. 4c**). Within the HIP, 21 miRNAs (13 upregulated and 8 downregulated) and 15 paired mRNAs (9 upregulated and 6 downregulated) could regulate four pathways (*G-Protein Coupled Receptor Signaling, CREB Signaling in Neurons, Synaptogenesis Signaling*, and *Axonal Guidance Signaling*) (**Fig. 4d**). Moreover, within the NAc, two upregulated miRNAs and two paired down-regulated mRNAs could regulate one pathway (i.e., *CREB Signaling in Neurons*) (**Fig. 5a**). Within the PFC, 19 miRNAs (14 upregulated and 5 downregulated) and 22 paired mRNAs (5 upregulated and 17 downregulated) could regulate four pathways (*IL-8 Signaling, Axonal Guidance Signaling, CREB Signaling in Neurons, and G-Protein Coupled Receptor Signaling*) (**Fig. 5b**). Within the PUT, four miRNAs (3 upregulated and 1 downregulated) and four paired mRNAs (2 upregulated and 2 downregulated) could regulate two pathways (*Sirtuin Signaling* and *IL-8 Signaling*) (**Fig. 5c**). Within the VTA, nine miRNAs (2 upregulated and 7 downregulated) and 10 mRNAs (9 upregulated and 1 downregulated) could regulate five pathways (*Opioid Signaling, CREB signaling in Neurons, IL-8 Signaling, NRF2-mediated Oxidative Stress Response*, and *Gap Junction Signaling*) (**Fig. 5d**). Most of the above AUD-related pathways could also be regulated by differentially expressed (absolute FC>1.3 & *P*<0.05) and negatively correlated miRNA-mRNA pairs identified in six of the above eight brain regions of AUD subjects (the Set 2 sample) (**Supplementary Fig. S13** and **Fig. S14**).

**Fig. 4.**
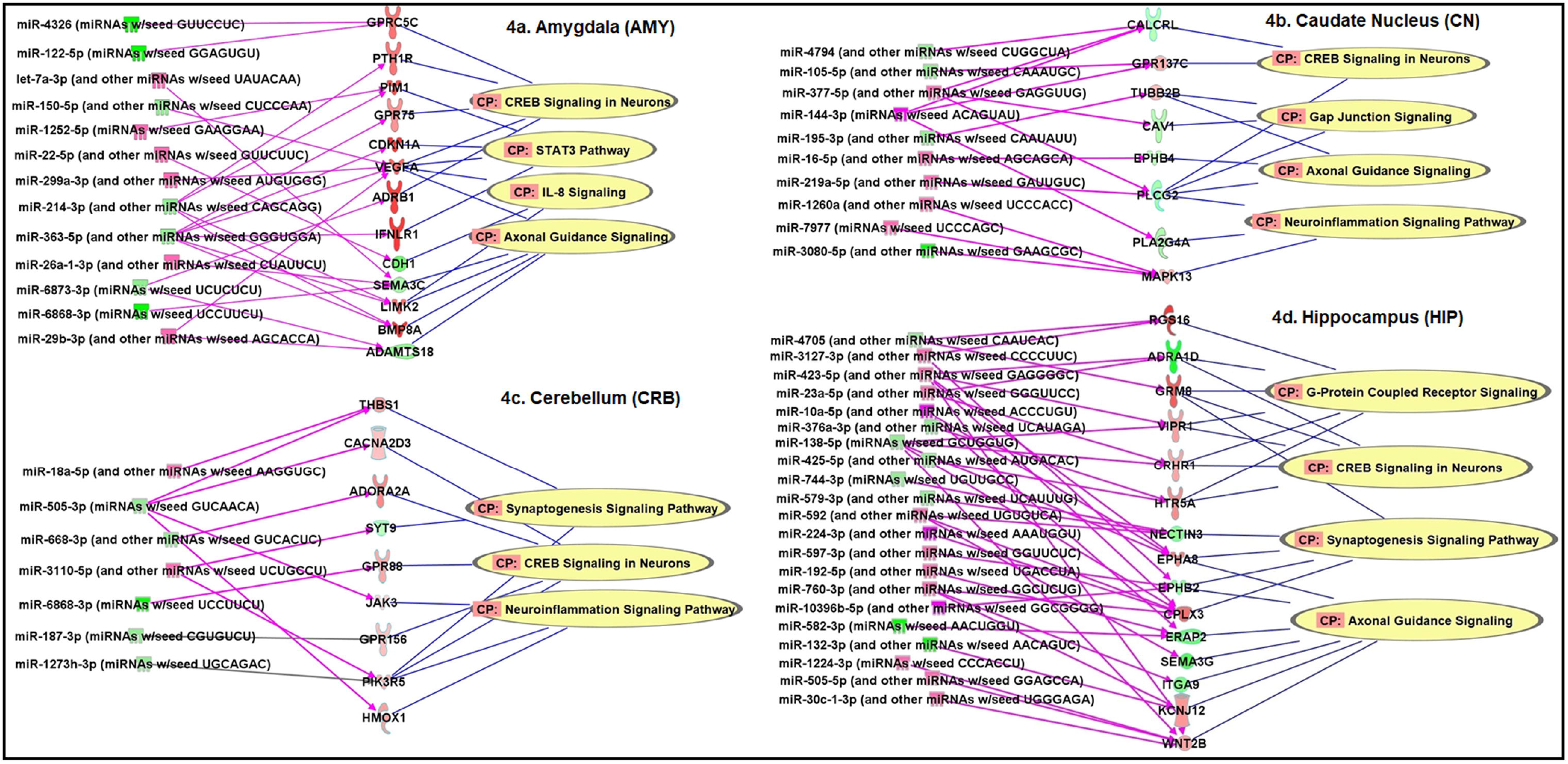
AUD-associated miRNA-mRNA regulatory networks in the amygdala (AMY), the caudate nucleus (CN), the cerebellum (CRB), and the hippocampus (HIP) of subjects with alcohol use disorder (AUD) (the Set 1 sample). CP: Canonical pathways potentially regulated by differentially expressed [absolute fold-change (FC) > 1.3 & *P* < 0.05] and negatively correlated miRNA-mRNA pairs identified in each brain region were defined using the Ingenuity Pathway Analysis (IPA) miRNA Target Filter function.

**Fig. 5.**
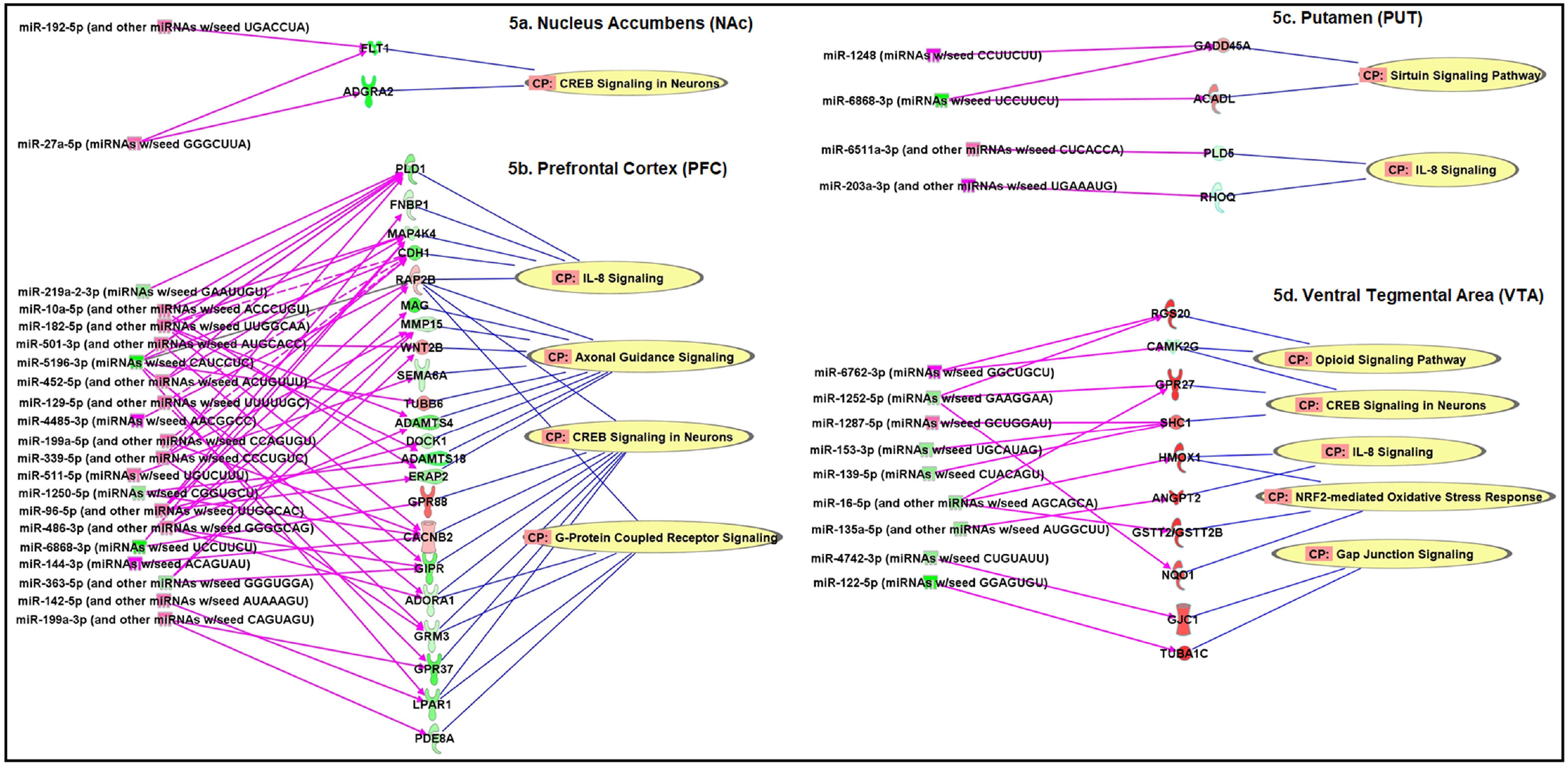
AUD-associated miRNA-mRNA regulatory networks in the nucleus accumbens (NAc), the prefrontal cortex (PFC), the putamen (PUT), and the ventral tegmental area (VTA) of subjects with alcohol use disorder (AUD) (the Set 1 sample). CP: Canonical pathways potentially regulated by differentially expressed [absolute fold-change (FC) > 1.3 & *P* < 0.05] and negatively correlated miRNA-mRNA pairs identified in each brain region were defined using the Ingenuity Pathway Analysis (IPA) miRNA Target Filter function.

The analysis of miRNA-mRNA regulatory networks using differentially expressed (absolute FC>1.3 & *P*<0.05) and negatively correlated miRNA-mRNA pairs identified in ethanol-exposed hESC-derived cortical interneurons indicated that four AUD-relevant pathways (*Sirtuin Signaling, Opioid Signaling, NRF2-mediated Oxidative Stress Response*, and *IL-1 Signaling*) could be regulated by nine miRNAs (3 upregulated and 6 downregulated) and six paired mRNAs (1 upregulated and 5 downregulated) (**Supplementary Fig. S15**). In total, 17 canonical pathways potentially regulated by differentially expressed and negatively correlated miRNA-mRNA pairs were identified in eight brain region of AUD subjects and ethanol-exposed hESC-derived cortical interneurons (**Supplementary Table S8**). The top three pathways potentially regulated by AUD-associated and negatively correlated miRNA-mRNA pairs in multiple brain regions included *CREB Signaling in Neurons, IL-8 Signaling*, and *Axonal Guidance Signaling*. Additionally, three AUD-associated miRNA-mRNA regulatory pathways (*Opioid Signaling Pathway, NRF-mediated Oxidative Stress Response*, and *Sirtuin Signaling*) were confirmed by the *in vitro* cellular model study (**Supplementary Table S8)**.

## Discussion

In this study, we observed miRNA and mRNA transcriptomic changes in multiple reward-related or alcohol-responsive brain regions of AUD subjects. We also discovered that miRNA-mRNA interactions in specific brain regions potentially contributed to the biological pathways important for AUD risk. Through the *in vitro* cellular model study, we validated that alcohol exposure could alter miRNA and mRNA expression profiles and miRNA-mRNA regulatory networks. To our knowledge, this is the first study that investigated the relationship of AUD and brain miRNA-mRNA regulatory networks.

First, RNA-seq and microarray analyses of transcriptomic changes in multiple brain regions of AUD subjects suggest that several cortical and subcortical regions (or components of the reward circuit) are essential for the rewarding effect of alcohol. We observed miRNA and mRNA transcriptomic changes in eight reward-related or alcohol-responsive brain regions of AUD subjects. The reason that we chose these eight brain regions for this study is that they participate in brain functions such as motivation, memory, and pleasure as well as balance and locomotion [39-41]. Certainly, we cannot exclude the possibility that other brain regions also mediate the rewarding effect of ethanol or be in involved in AUD-related pathways.

Second, a brain region with more AUD-associated miRNAs and mRNAs may be more responsive to alcohol stimulation or play a more important role in alcohol-induced neuroadaptations. As shown in **Figure 1**, the PFC had the largest number of AUD-associated miRNAs, and there were more upregulated than downregulated miRNAs in the PFC. Correspondingly, more mRNAs were significantly downregulated in the PFC of AUD subjects than in other brain regions of AUD subjects (**Figure 2**). Given the role of the PFC in higher cognitive functions, alcohol-induced expression changes of miRNAs and their target mRNAs in the PFC may lead to cognitive deficits and compromised working memory. Moreover, differentially expressed miRNAs and mRNAs were also observed in seven other brain regions of AUD subjects, and some AUD-associated miRNAs and mRNAs were shared among multiple brain regions of AUD subjects (**Fig. 1** and **Fig. 2**). These findings provided insight into the coordinated role of multiple brain regions in AUD development and also suggested coordinated expression changes of miRNAs and mRNAs in the brains of AUD subjects.

Third, the findings that AUD-associated brain miRNAs potentially target genes involved in addiction-linked pathways suggest that these miRNAs play a critical role in AUD development. Through miRNA target gene prediction and pathway enrichment analyses by DIANA-mirPath, we found that the majority of the 19 differentially expressed miRNAs (**Supplementary Table S2**) identified in one or more of the eight brain regions could target coding genes (or mRNAs) that participate in neurobiological processes of drug reward or addiction (**Fig. 3**). Among the top 14 pathways, four were related to drug addiction (*Morphine Addition, Retrograde Endocannabinoid Signaling, Cocaine Addiction*, and *Amphetamine Addiction*) and two were related to synaptic functions (*GABAergic Synapse* and *Glutamatergic Synapse*). Although alcohol and drugs of abuse (e.g., morphine and cocaine) possess diverse neuropharmacological potentials, their reinforcing effects are mediated by common pathways (such as dopaminergic and glutamatergic pathways) *via* the activation of the mesocorticolimbic system that are mainly comprised of the AMY, the NAc, the PFC, and the VTA [42]. That is to say, the above pathways for drug addiction or synaptic function can also mediate the rewarding effect of alcohol or are essential for neuroadaptive processes triggered by alcohol. Accordingly, AUD-associated miRNAs identified in the above eight brain regions are expected to regulate the expression of genes that are important for alcohol-induced neuroadaptations.

Additionally, the present study provided evidence that brain miRNA-mRNA regulatory networks consisting of dysregulated and negatively correlated miRNA-mRNA pairs contribute to the risk of AUD. We identified at least 17 canonical pathways that were likely influenced by dysregulated and negatively correlated miRNA-mRNA pairs in the brains of AUD subjects (**Supplementary Table S8**). The top three pathways potentially regulated by dysregulated and negatively correlated miRNA-mRNA pairs in multiple brain regions of AUD subjects included *CREB Signaling in Neurons, IL-8 Signaling*, and *Axonal Guidance Signaling*. The *CREB Signaling* was found to be a central amygdaloid signaling pathway involved in high anxiety-like and excessive alcohol drinking behaviors [43]. We found that the *CREB Signaling* pathway could be regulated by dysregulated and negatively correlated miRNA-miRNA pairs in seven of the eight brain regions (except PUT) of AUD subjects (**Fig. 4** and **Fig. 5**). Regarding the relationship of the *IL-8 Signaling* pathway and AUD, there is emerging evidence that alcohol use can stimulate immune cells to secrete peripheral pro- and anti-inflammatory cytokines (such as IL-8) [44, 45], thus supporting the role of the immune system in the pathophysiology of AUD. We observed that the *IL-8 Signaling* pathway could be regulated by dysregulated and negatively correlated miRNA-miRNA pairs in four (AMY, PFC, PUT, and VTA) of the eight brain regions of AUD subjects (**Fig. 4** and **Fig. 5**). The *Axon Guidance Signaling* pathway can regulate axon guidance, synaptogenesis, and cell migration. Studies have shown that ethanol disrupted axon outgrowth by influencing the *Axon Guidance Signaling* pathway [46]. We noticed that the *Axon Guidance Signaling* pathway could be regulated by dysregulated and negatively correlated miRNA-miRNA pairs in four (AMY, CN, HIP, and PFC) of the eight brain regions of AUD subjects (**Fig. 4** and **Fig. 5**). These three top pathways were validated in the Set 2 brain tissue sample by the network analysis of dysregulated and negatively correlated miRNA-mRNA pairs in six of the eight brain regions of AUD subjects (**Supplementary Fig. S13** and **S14**). Although these three top pathways were not found to be regulated by differentially expressed and negatively correlated miRNA-mRNA pairs identified in ethanol-exposed hESC-derived cortical interneurons, four other pathways [*Sirtuin Signaling, Opioid Signaling, NRF2-mediated Oxidative Stress Response*, and *interleukin-1* (*IL-1*) *Signaling*] were uncovered (**Supplementary Table S15**). Except the *IL-1 Signaling* pathway, three other pathways were also identified by the analysis of Set 1 and Set 2 samples. Similar to the *IL-8 Signaling* pathway, the *IL-1 Signaling* pathway can also regulate immune response or inflammation caused by alcohol [47]. Therefore, multiple addiction-linked pathways influenced by AUD-associated miRNA-mRNA regulatory networks could contribute to the occurrence of AUD.

Some limitations of this study should be noted. First, bulk RNA-seq cannot evaluate the functional relevance of miRNA-mRNA pairing at the cellular level. Since RNA samples for the transcriptome analysis were extracted from homogenized brain tissues, AUD-associated miRNA and mRNA expression changes may not occur in the same type of cells. To identified AUD-associated and cell type-specific miRNA-mRNA pairs, single-cell (or nucleus) RNA-seq can be applied to map miRNA and mRNA transcriptomes at the individual cell level. Second, the functional role of AUD-associated miRNA-mRNA networks in regulating neuronal function was not investigated. We only predicted by bioinformatics programs or based on published studies that a number of AUD-related pathways were regulated by AUD-associated and negatively correlated miRNA-mRNAs pairs. Animal model studies can be conducted to determine the influence of miRNA-mRNA interactions on neuronal function and addiction-related behaviors. Third, the transcriptome analysis of postmortem brain tissues cannot determine whether the dysregulation of brain miRNAs and mRNAs was due to pre-existing vulnerability factors (such as genetic variants and/or environmental insults) or long-term alcohol consumption. We intended to verify AUD-associated brain miRNA and mRNA changes using ethanol-exposed hESC-derived cortical interneurons as models. However, not many AUD-associated brain miRNAs and mRNAs were validated by the *in vitro* cellular model study. To confirm whether AUD-associated brain miRNA and mRNA expression changes were indeed due to alcohol use and occur in a certain type of brain neuronal or glial cells, we could use controlled animal model studies and single-cell (or nucleus) RNA-seq. Additionally, we did not analyze other types of AUD-associated noncoding RNAs, such as long noncoding RNAs (lncRNAs). In the follow-up study, we will further analyze AUD-associated miRNA-mRNA-lncRNA regulatory networks.

In conclusion, the concerted expression changes of brain miRNAs and their target mRNAs as well as the interaction of them may govern alcohol-induced neuroplasticity, thus contributing to the development of AUD. To understand the mechanisms of the transition of alcohol use to abuse or dependence, the spatial and temporal expression of brain miRNAs and their target mRNAs need to be investigated.

## Data Availability

The transcriptome data has been submitted to
the Gene Expression Omnibus database.

https://www.ncbi.nlm.nih.gov/geo/

## Acknowledgements

This work was supported by the National Institute on Alcohol Abuse and Alcoholism grant R01AA025080 (HZ) and R21AA023068 (HZ). The authors are grateful to the New South Wales Tissue Resource Centre (NSWTRC) at The University of Sydney, Australia for providing alcoholic and control brain tissues for this study. The NSWTRC is supported by the University of Sydney, the National Health and Medical Research Council of Australia, and the National Institute on Alcohol Abuse and Alcoholism. We also thank the deceased subjects’ next of kin for providing informed written consent for the studies. Additionally, the authors wish to thank Christopher Castaldi and Bryan Pasqualucci at the Yale Center for Genome Analysis (YCGA) as well as Mei Zhong and Ee-Chun Cheng at the Yale Stem Cell Center (YSCC) for helping with the RNA sequencing experiments.

## Conflict of Interest

The authors declare no conflict of interest.

Supplementary information is available at MP’s website.

## Notes

### Competing Interest Statement

The authors have declared no competing interest.

### Funding Statement

This work was supported by the National Institute on Alcohol Abuse and Alcoholism grant
R01AA025080 and R21AA023068.

### Author Declarations

The Institutional Review Board (IRB) of Boston University has reviewed the study "Brain microRNA-mRNA regulatory networks and alcohol use disorder" (IRB number: H-36701). Decision: the ethical approval was waived since this is not a human subjects research.

