## Supplementary materials for "Exploration of alcohol use disorder-associated brain miRNA-mRNA regulatory networks"

**Fig. S1** Principal component analysis (PCA) of miRNA-seq (a) and rRNA depletion RNA-seq (b) data of the 192 selected Set 1 RNA samples.
**Fig. S2** Box plotting of the Affymetrix miRNA (a) and mRNA (b) transcriptome data of the 96 selected Set 2 RNA samples.
**Fig. S3** Morphology and immunostaining of cortical interneurons derived from H1 human embryonic stem cells (hESCs).
**Fig. S4.** Comparison of the morphology of hESC-derived cortical interneurons with or without ethanol exposure for 7 days.
**Fig. S5** Box plotting of miRNA (a) and mRNA (b) transcriptomes of hESC-derived cortical interneurons with or without ethanol exposure.
**Fig. S6** Volcano plots displaying differentially expressed miRNAs in six regions of postmortem brains of subjects with alcohol use disorder (AUD) (the Set 2 sample).
**Fig. S7** Volcano plot highlighting differentially expressed miRNAs in hESC-derived cortical interneurons exposed to ethanol for 7 days.
**Fig. S8** Venn diagrams showing the number of differentially expressed miRNAs ($P$<0.05) shared between eight brain regions of AUD subjects (Set 1 and Set 2) and ethanol-exposed hESC-derived cortical interneurons.
**Fig. S9** Volcano plots displaying differentially expressed mRNAs in six regions of postmortem brains of subjects with alcohol use disorder (AUD) (the Set 2 sample).
**Fig. S10** Volcano plot displaying differentially expressed mRNAs in hESC-derived cortical interneurons exposed to ethanol for 7 days.
**Fig. S11** Venn diagrams showing the number of differentially expressed mRNAs ($P$ < 0.05) shared between eight brain regions of AUD subjects (Set 1 and Set 2) and ethanol-exposed hESC-derived cortical interneurons.
**Fig. S12** Gene ontology (GO) analysis of 97 differentially expressed mRNAs (absolute FC > 2.0 & $P$ < 0.001) identified in one or more of the eight brain regions of AUD subjects (the Set 1 sample).
**Fig. S13** AUD-associated miRNA-mRNA regulatory networks in the amygdala (AMY), the caudate nucleus (CN), the cerebellum (CRB) of subjects with alcohol use disorder (AUD) (the Set 2 sample).
**Fig. S14** AUD-associated miRNA-mRNA regulatory networks in the hippocampus (HIPPO), the prefrontal cortex (PFC), and the putamen (PUT) of subjects with alcohol use disorder (AUD) (the Set 2 sample).
**Fig. S15** miRNA-mRNA regulatory networks in ethanol-exposed human embryonic stem cell (hESC)-derived cortical interneurons.

**Table S1** Demographic details of brain tissue samples.
**Table S2** Small RNA-seq analysis identified miRNAs with differential expression in eight brain regions of AUD subjects (Set 1).
**Table S3** Microarray analysis identified miRNAs with differential expression in six brain regions of AUD subjects (Set 2).
**Table S4** Small RNA-seq analysis identified miRNAs with differential expression in hESC-derived cortical interneurons with ethanol exposure.
**Table S5** rRNA depletion RNA-seq analysis identified mRNAs with differential expression in eight brain regions of AUD subjects (Set 1).
**Table S6** Microarray analysis identified mRNAs with differential expression in six brain regions of AUD subjects (Set 2).
**Table S7** mRNA-seq analysis identified mRNAs with differential expression in ethanol-exposed hESC-derived cortical interneurons.
**Table S8** Canonical pathways potentially regulated by differentially expressed and negatively correlated miRNA-mRNA pairs.

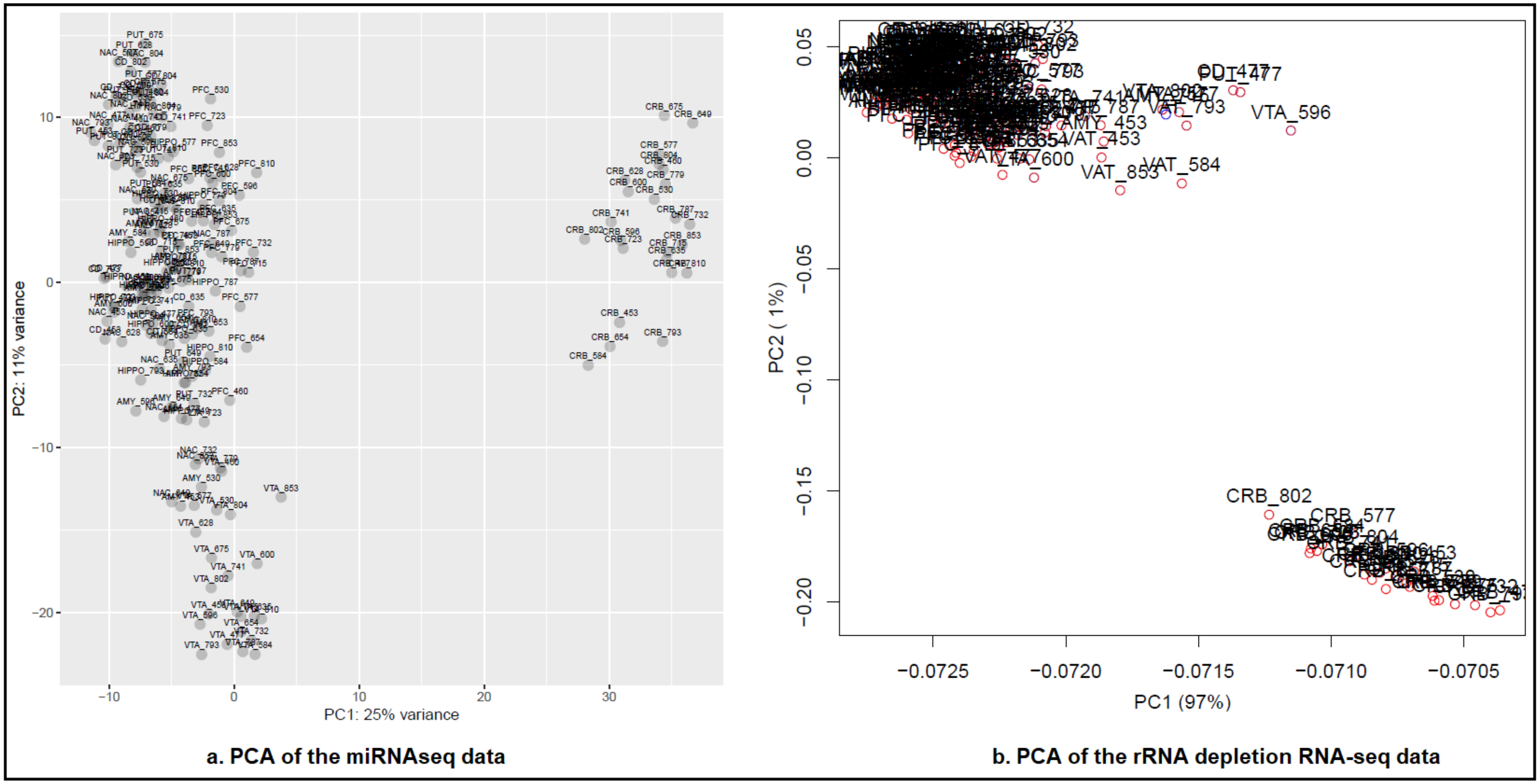


**Fig. S1** Principal component analysis (PCA) of miRNA-seq (a) and rRNA depletion RNA-seq (b) data of the 192 selected Set 1 RNA samples.

AMY: Amygdala; CN: Caudate Nucleus; CRB: Cerebellum; HIPPO: Hippocampus; NAc: Nucleus Accumbens; PFC: Prefrontal Cortex; PUT: Putamen; and VTA: Ventral Tegmental Area.

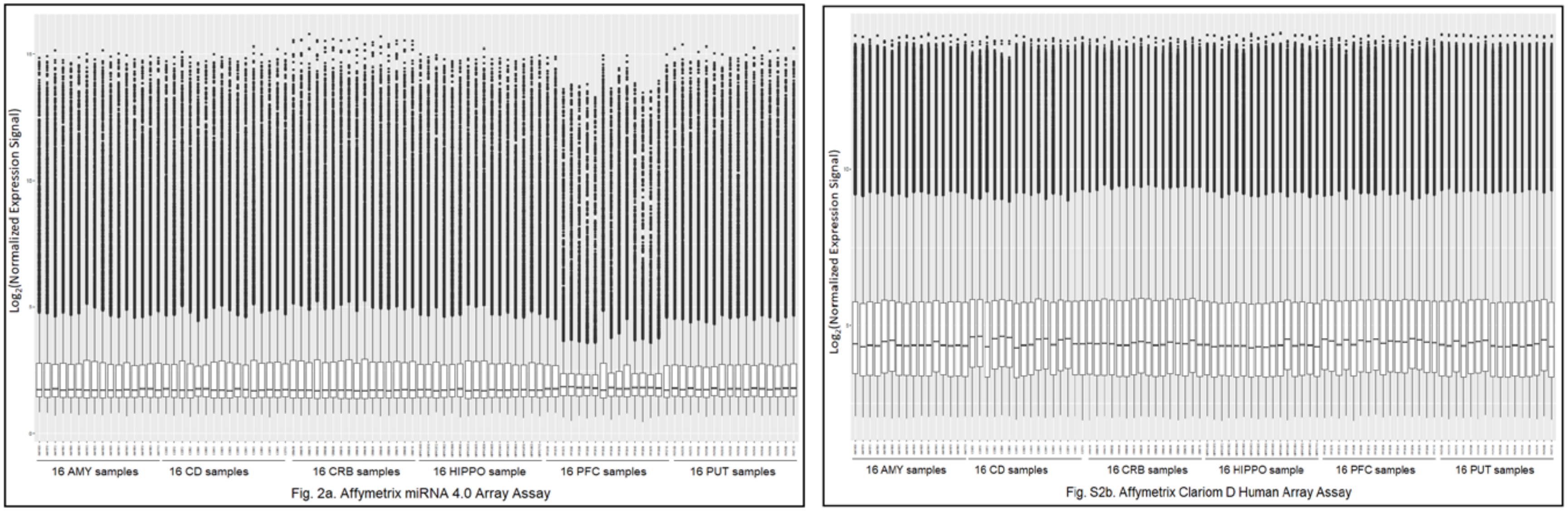


**Fig. S2** Box plotting of the Affymetrix miRNA (a) and mRNA (b) transcriptome data of the 96 selected Set 2 RNA samples.
a: Box plotting of $log_2$ expression signals of miRNAs mapped by Affymetrix miRNA 4.0 Array assay.
b: Box plotting of $log_2$ expression signals of mRNA transcripts mapped by Affymetrix Clariom D Human Array assay.

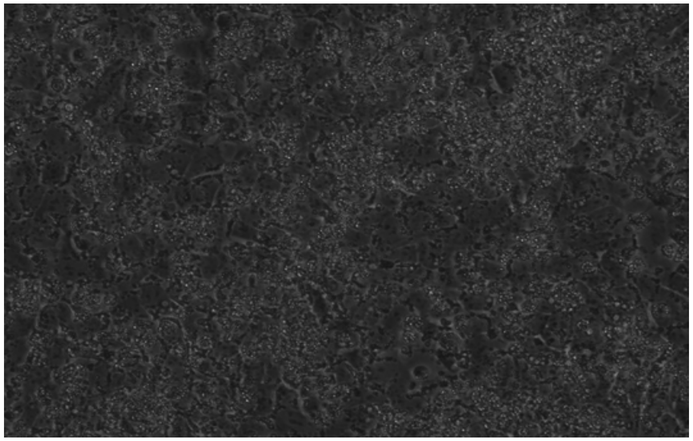

a. Day 11 - Start patterning

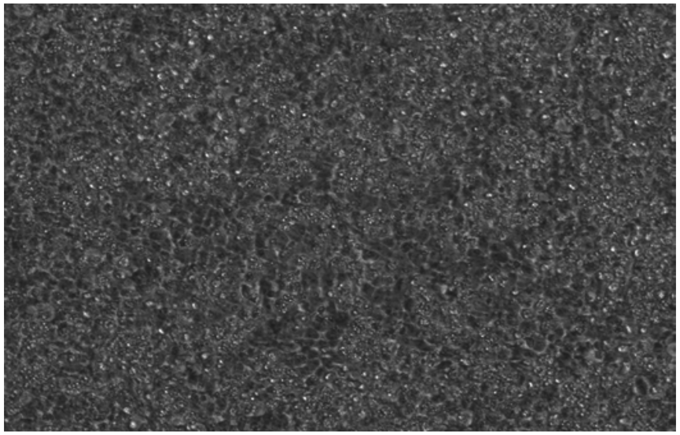

b. Day 19 - Start maturing

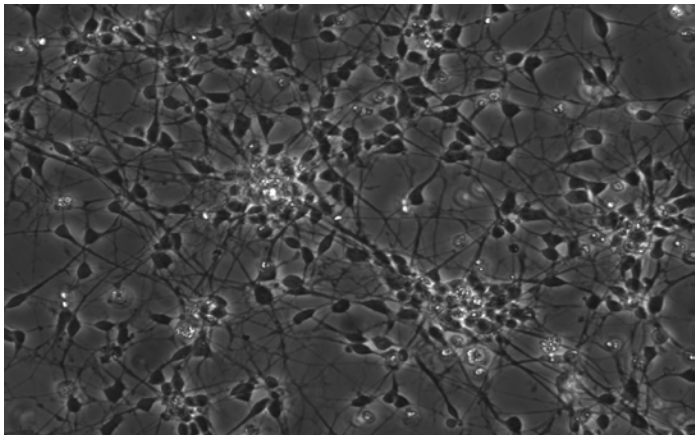

c. Day 33 - 2-week maturation

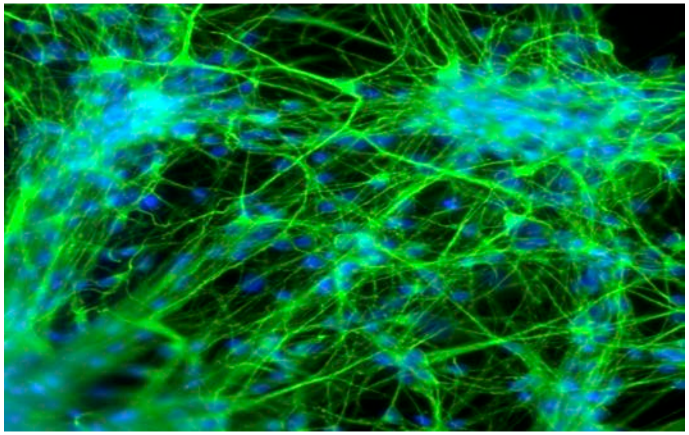

d. Day 62 - Immunostaining

**Fig. S3** Morphology and immunostaining of cortical interneurons derived from H1 human embryonic stem cells (hESCs).
a. Morphology of cells starting patterning at Day 11.
b. Morphology of cells starting maturing at Day 19.
c. Morphology of cells after a 2-week maturation at Day 33.
d. Immunostaining of cells at Day 62 using antibodies reacting with beta III tubulin, a biomarker of GABAergic neurons.

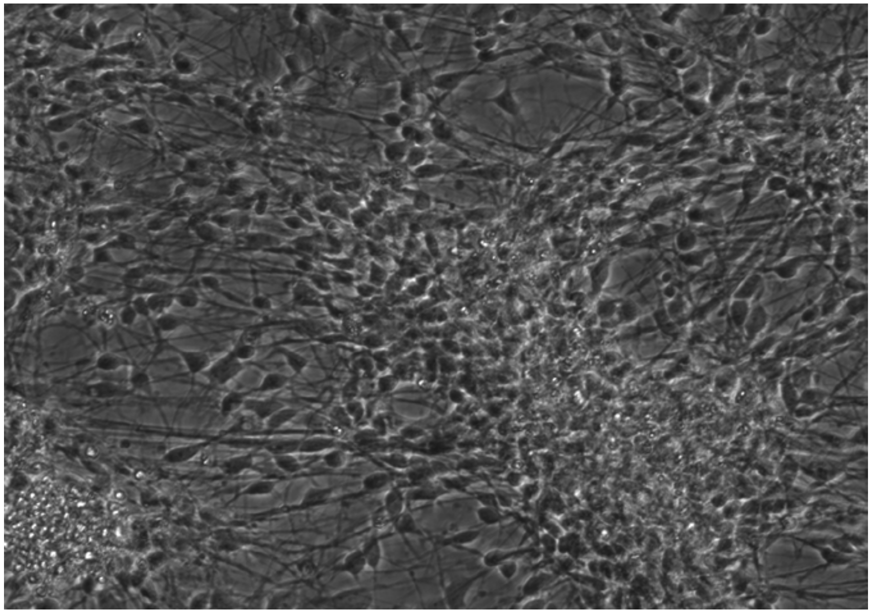

A 7-day culture - No ethanol

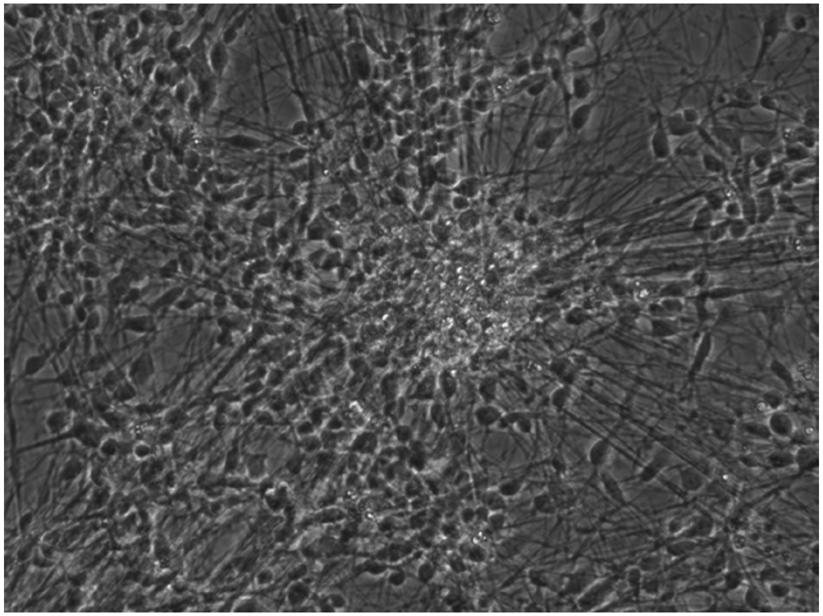

A 7-day culture - with ethanol

**Fig. S4.** Comparison of the morphology of hESC-derived cortical interneurons with or without ethanol exposure for 7 days.

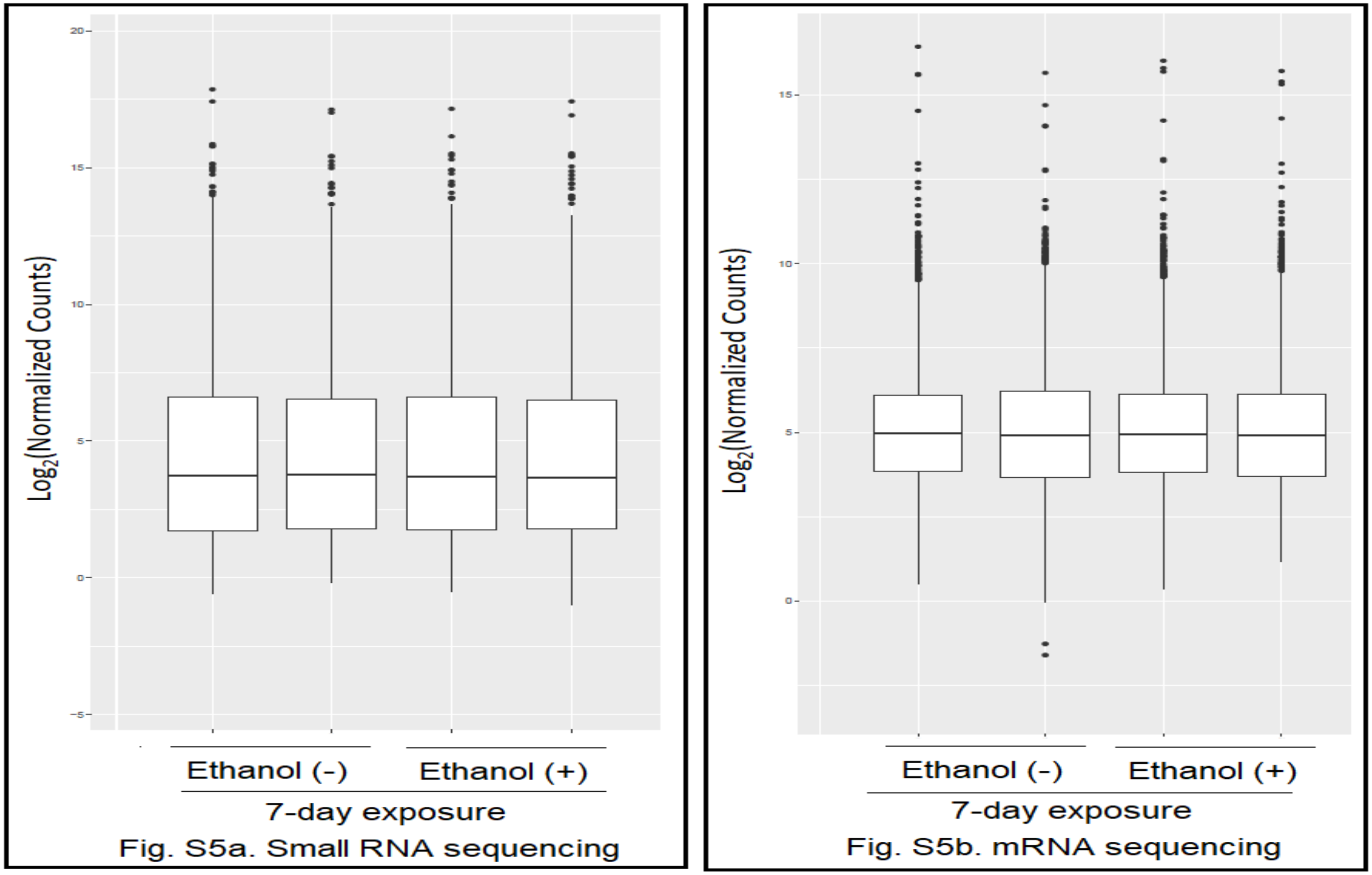


**Fig. S5** Box plotting of miRNA and mRNA transcriptomes of hESC-derived cortical interneurons with or without ethanol exposure.
a: Box plotting of $\log_2$ normalized counts of mature miRNAs mapped by small RNA sequencing.
b: Box plotting of $\log_2$ normalized counts of mRNA transcripts mapped by mRNA sequencing.

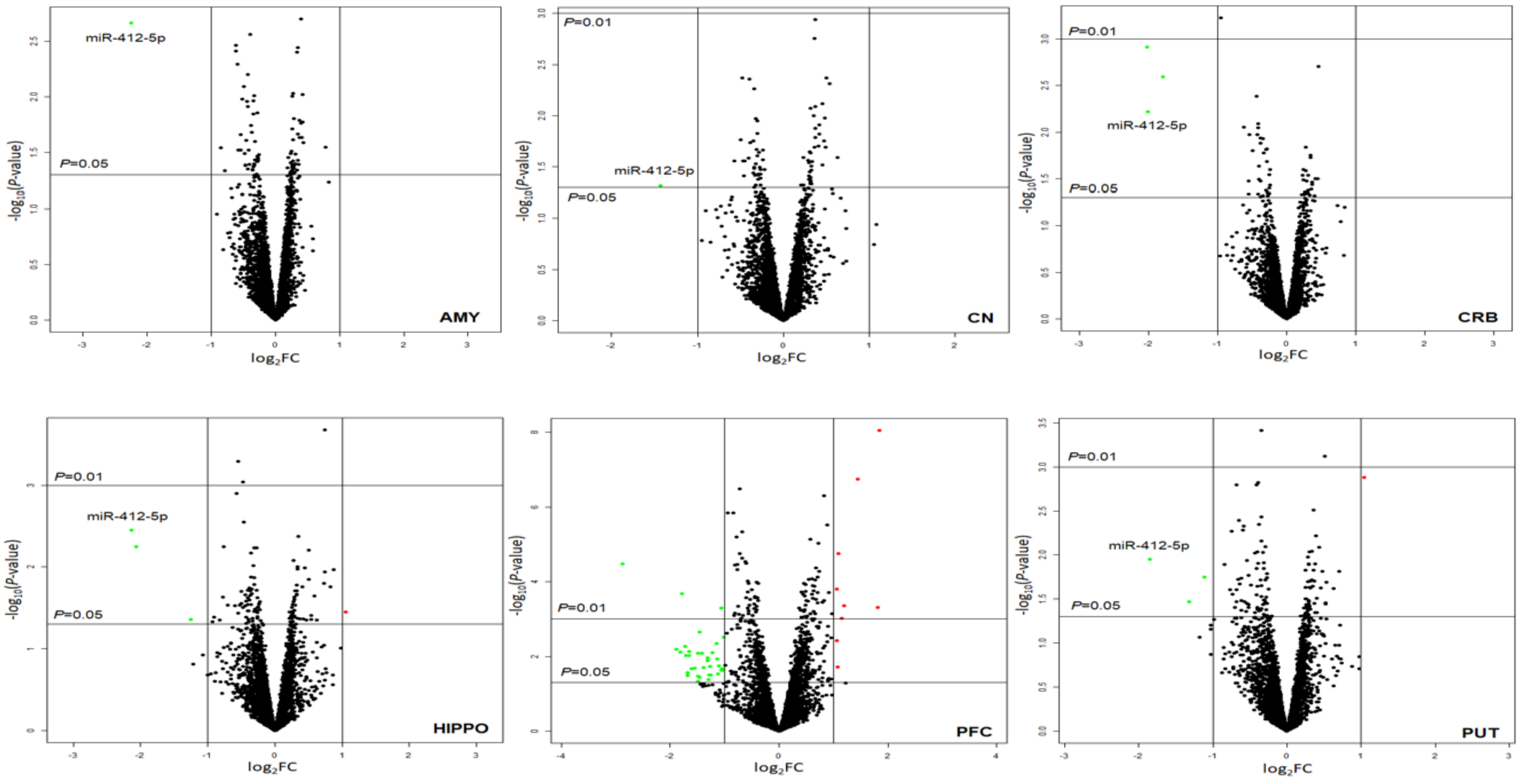


**Fig. S6** Volcano plots displaying differentially expressed miRNAs in six regions of postmortem brains of subjects with alcohol use disorder (AUD) (the Set 2 sample).

The vertical axis (y-axis) corresponds to the negative $\log_{10}$ of the *P*-value, and the horizontal axis (x-axis) displays the $\log_2$ of fold changes (FC). The red dots represent up-regulated miRNAs ($\log_2$FC > 1.0 & $P < 0.05$) and the green dots represent downregulated miRNAs ($\log_2$FC < -1.0 & $P < 0.05$). The horizontal line shows the *P*-value cutoff ($P = 0.05$ or 0.01) with points above the line having the *P*-value < 0.05 or 0.01 and points below the line having the *P*-value > 0.05 or 0.01. The two vertical lines indicate 2-fold changes. miR-412-5p was downregulated in five of the six brain regions. AMY: Amygdala; CN: Caudate Nucleus; CRB: Cerebellum; HIPPO: Hippocampus; PFC: Prefrontal Cortex; PUT: Putamen.

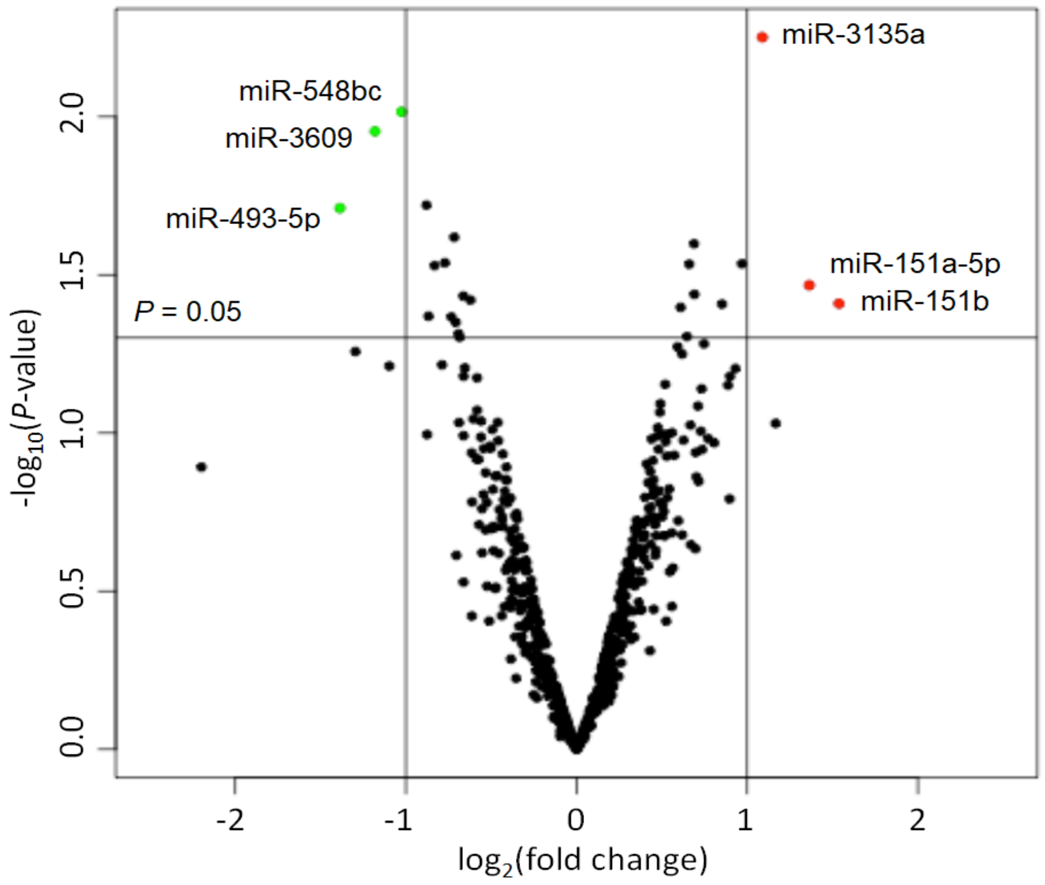


**Fig. S7** Volcano plot highlighting differentially expressed miRNAs in hESC-derived cortical interneurons exposed to ethanol for 7 days.

The vertical axis (y-axis) corresponds to the negative $\log_{10}$ of the *P*-value, and the horizontal axis (x-axis) displays the $\log_2$ of fold changes (FC). The red dots represent up-regulated miRNAs ($\log_2$FC > 1.0 & $P < 0.05$), and the green dots represent downregulated miRNAs ($\log_2$FC < -1.0 & $P < 0.05$). The horizontal line shows the *P*-value cutoff ($P = 0.05$) with points above the line having the *P*-value < 0.05 and points below the line having the *P*-value > 0.05. The two vertical lines indicate 2-fold changes.

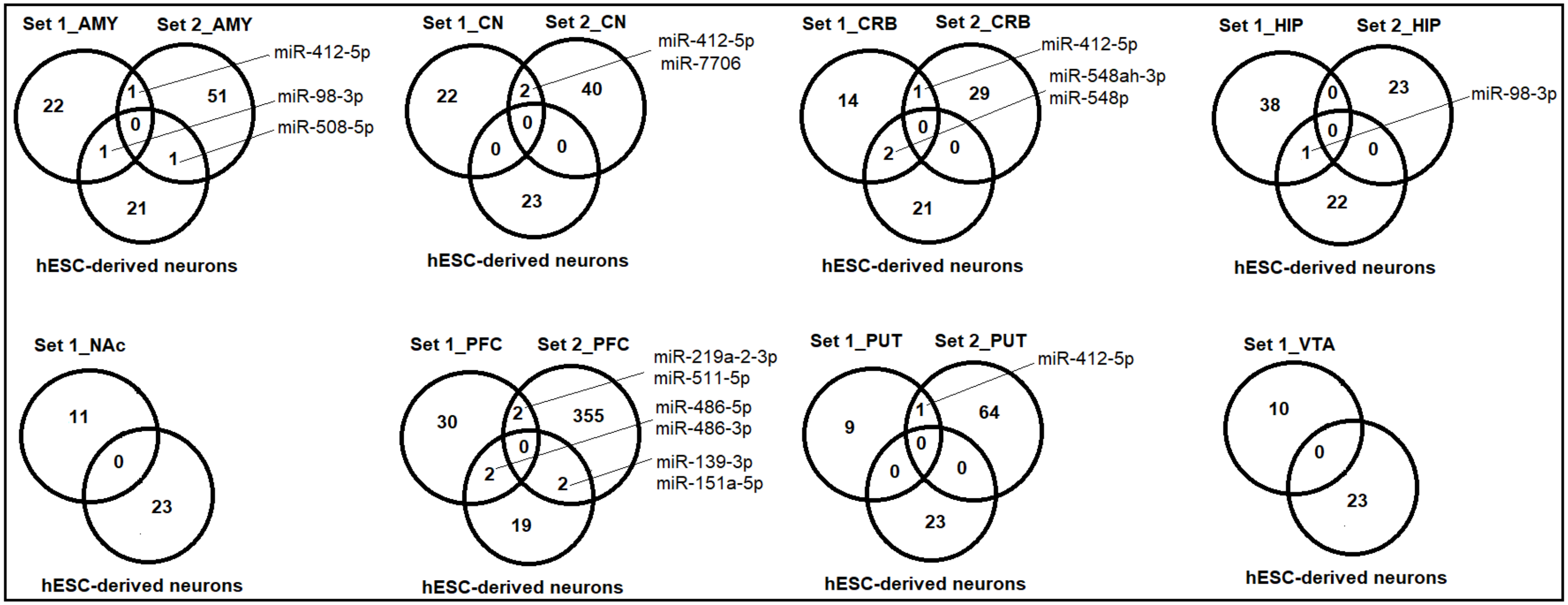


**Fig. S8** Venn diagrams showing the number of differentially expressed miRNAs ($P$<0.05) shared between eight brain regions of AUD subjects (Set 1 and Set 2) and ethanol-exposed hESC-derived cortical interneurons.
AMY: Amygdala; CN: Caudate Nucleus; CRB: Cerebellum; HIP: Hippocampus; NAc: Nucleus Accumbens; PFC: Prefrontal Cortex; PUT: Putamen; and VTA: Ventral Tegmental Area.

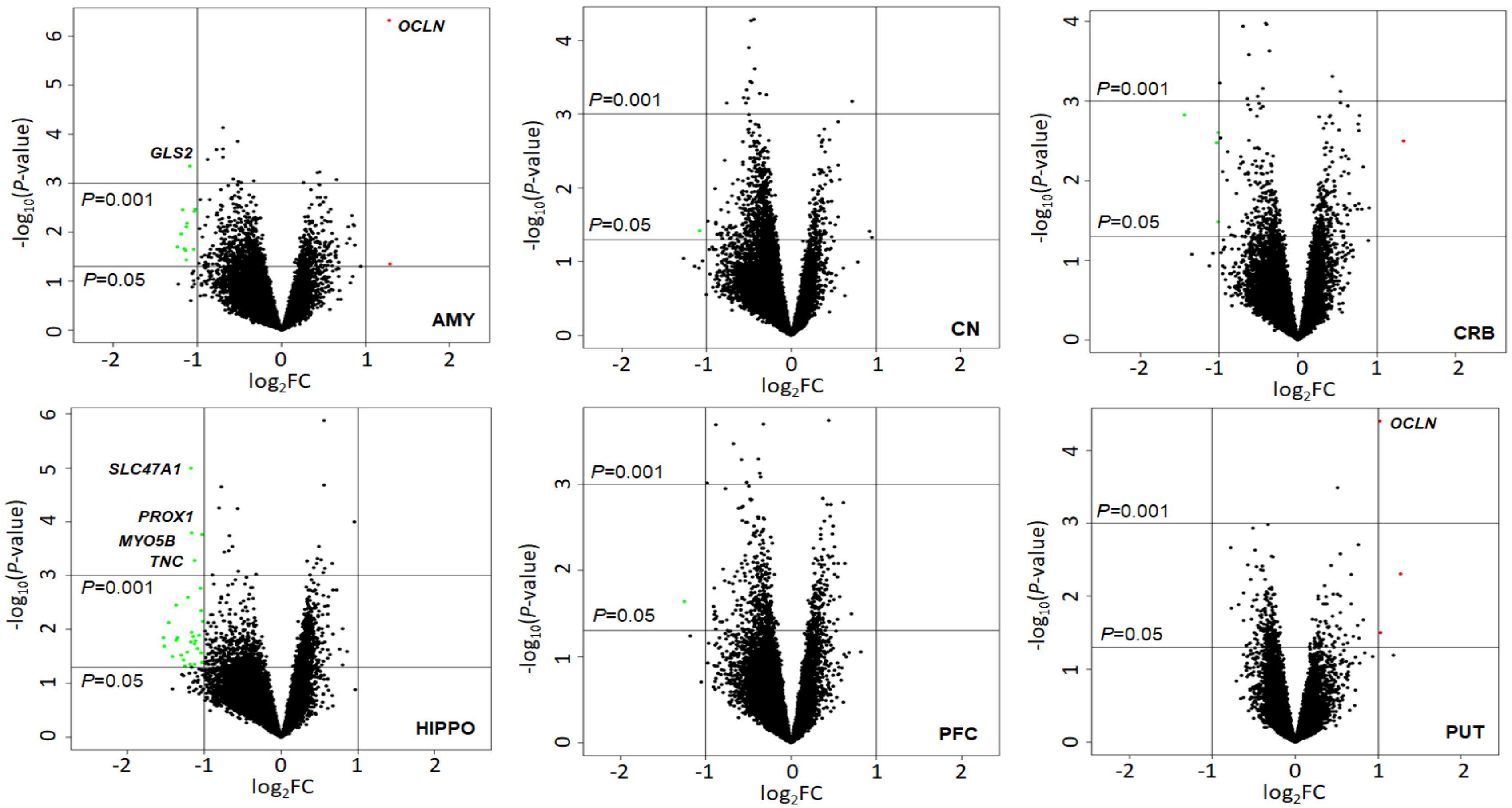


**Fig. S9** Volcano plots displaying differentially expressed mRNAs in six regions of postmortem brains of subjects with alcohol use disorder (AUD) (the Set 2 sample).

The vertical axis (y-axis) corresponds to the negative $\log_{10}$ of the *P*-value, and the horizontal axis (x-axis) displays the $\log_2$ of fold changes (FC). The red dots represent up-regulated miRNAs ($\log_2$FC > 1.0 & $P < 0.05$), and the green dots represent downregulated miRNAs ($\log_2$FC < -1.0 & $P < 0.05$). The horizontal line shows the *P*-value cutoff ($P = 0.05$ or 0.01) with points above the line having the *P*-value < 0.05 or 0.01 and points below the line having the *P*-value > 0.05 or 0.01. The two vertical lines indicate 2-fold changes. *OCLN* was upregulated and *GLS2*, *SLC47A1*, *PROX1*, *MYO5B*, and *TNC* were downregulated in multiple brain regions AUD subjects.
AMY: Amygdala; CN: Caudate Nucleus; CRB: Cerebellum; HIPPO: Hippocampus; PFC: Prefrontal Cortex; PUT: Putamen.

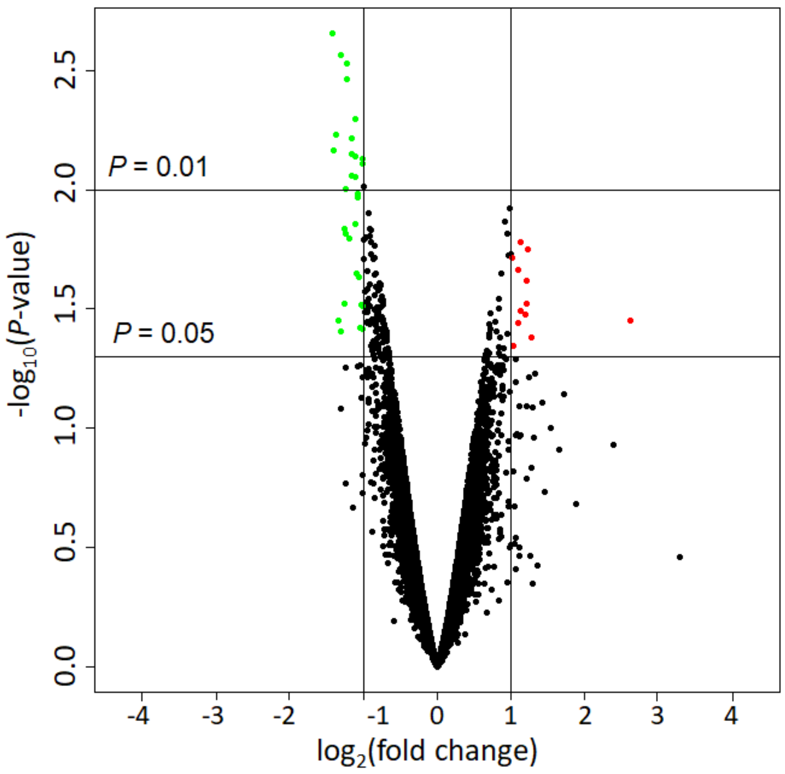


**Fig. S10** Volcano plot displaying differentially expressed mRNAs in hESC-derived cortical interneurons exposed to ethanol for 7 days.

The vertical axis (y-axis) corresponds to the negative $\log_{10}$ of the *P*-value, and the horizontal axis (x-axis) displays the $\log_2$ of fold changes (FC). The red dots represent up-regulated mRNAs ($\log_2$FC > 1.0 & $P < 0.05$) and the green dots represent downregulated miRNAs ($\log_2$FC < -1.0 & $P < 0.05$). The horizontal line shows the *P*-value cutoff ($P = 0.05$ or 0.01) with points above the line having the *P*-value < 0.05 or 0.01 and points below the line having the *P*-value > 0.05 or 0.01. The two vertical lines indicate 2-fold changes.

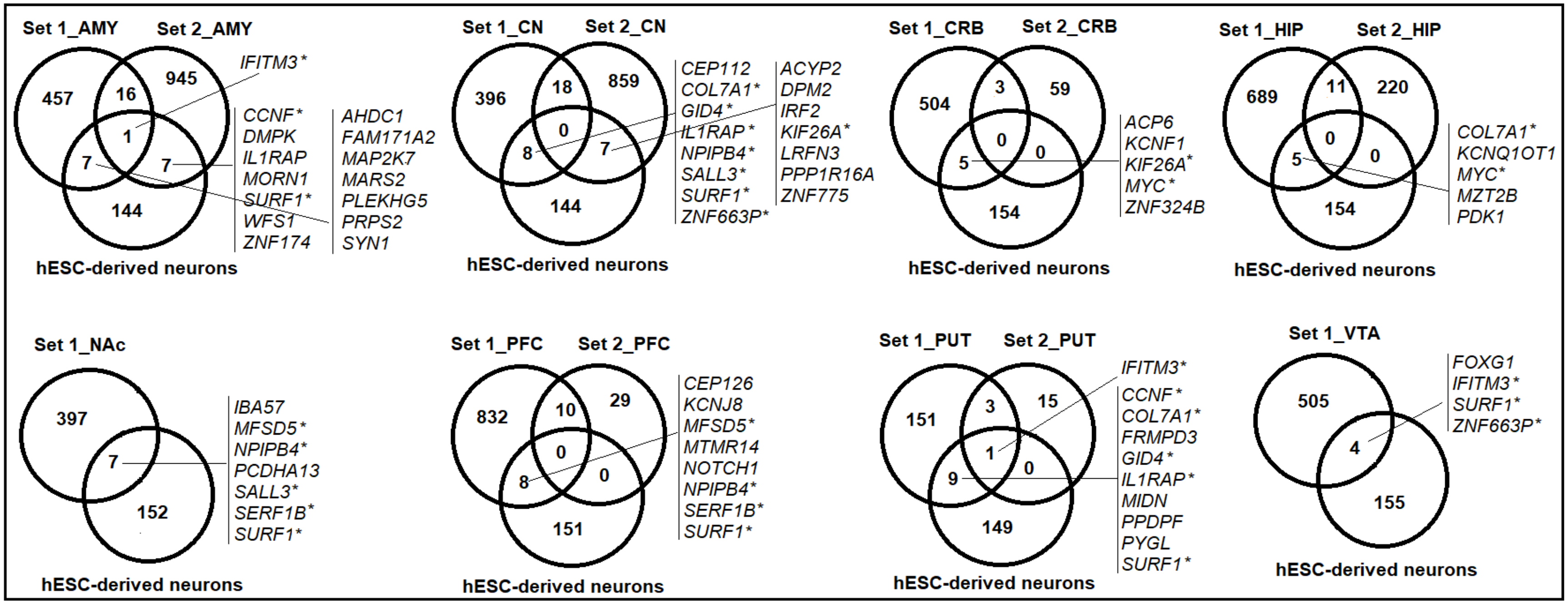


**Fig. S11** Venn diagrams showing the number of differentially expressed mRNAs ($P < 0.05$) shared between eight brain regions of AUD subjects (Set 1 and Set 2) and ethanol-exposed hESC-derived cortical interneurons.
AMY: Amygdala; CN: Caudate Nucleus; CRB: Cerebellum; HIP: Hippocampus; NAc: Nucleus Accumbens; PFC: Prefrontal Cortex; PUT: Putamen; and VTA: Ventral Tegmental Area.
*13 AUD-associated coding genes ($P < 0.05$) identified in multiple brain regions were found differentially expressed in ethanol-exposed hESC-derived cortical interneurons. They were *CCNF* (from AMY and PUT), *COL7A1* (from CN, HIP, and PUT), *GID4* (from CN and PUT), *IFITM3* (from AMY, PUT, and VTA), *IL1RAP* (from CN and PUT), *KIF26A* (from CN and CRB), *MFSD5* (from NAc and PFC), *MYC* (from CRB and HIP), *NPIPB4* (from CN, NAc, and PFC), *SALL3* (from CN and NAc), *SERF1B* (from NAc and PFC), *SURF1* (from AMY, CN, NAc, PFC, PUT, and VTA), and *ZNF663P* (from CN and VTA).

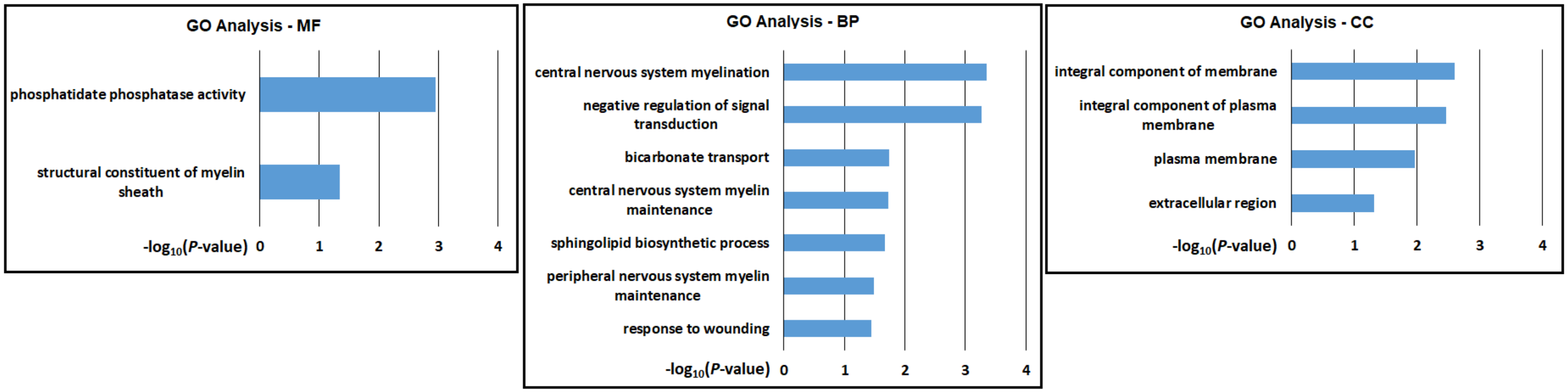


**Fig. S12** Gene ontology (GO) analysis of 97 differentially expressed mRNAs (absolute FC > 2.0 & $P < 0.001$) identified in one or more of the eight brain regions of AUD subjects (the Set 1 sample).
MF: molecular function; BP; biological process; and CC: cellular component. $-\log_{10}(P\text{-value})$ is the negative logarithm of *P*-value; a larger $-\log_{10}(P\text{-value})$ indicates a smaller *P*-value. GO terms with $-\log_{10}(P\text{-value}) > 1.3$ (or $P < 0.05$) were displayed.

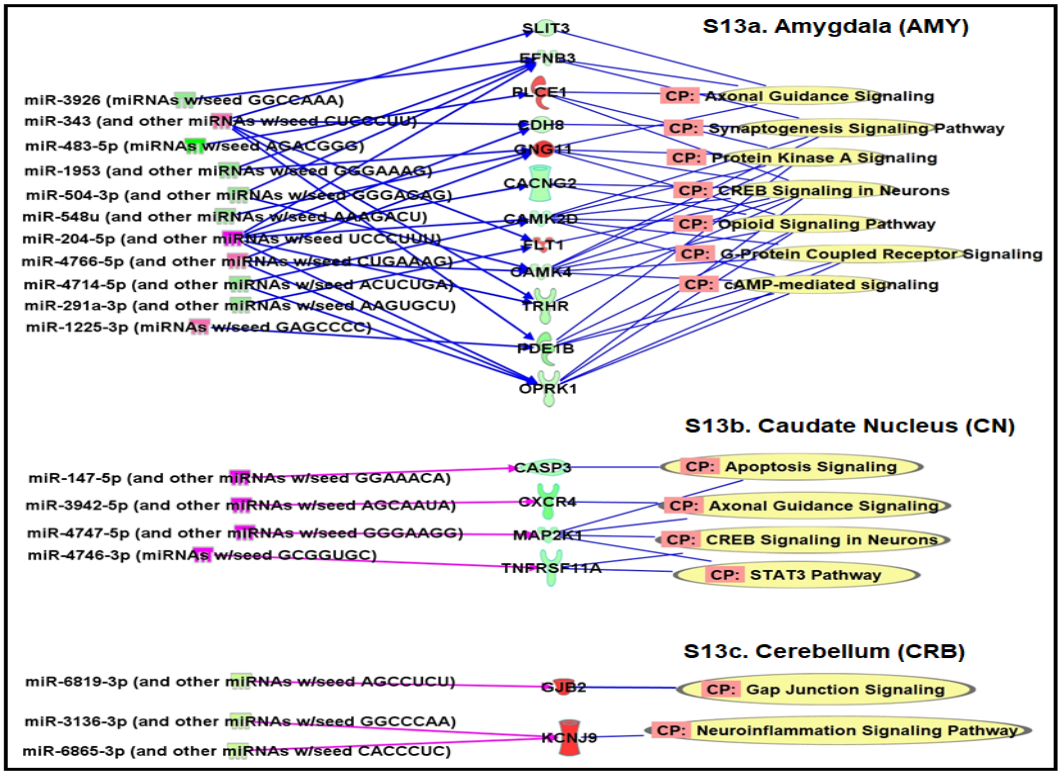


**Fig. S13** AUD-associated miRNA-mRNA regulatory networks in the amygdala (AMY), the caudate nucleus (CN), the cerebellum (CRB) of subjects with alcohol use disorder (AUD) (the Set 2 sample).

CP: Canonical pathways potentially regulated by differentially expressed [absolute fold-change (FC) > 1.3 & $P < 0.05$] and negatively correlated miRNA-mRNA pairs identified in each brain region were defined using the Ingenuity Pathway Analysis (IPA) miRNA Target Filter function. Within the amygdala (AMY), 11 miRNAs (4 upregulated and 7 downregulated) and 12 paired mRNAs (3 upregulated and 9 downregulated) could regulate seven pathways (*Axonal Guidance Signaling*, *Synaptogenesis Signaling*, *Protein Kinase A Signaling*, *CREB Signaling*, *Opioid Signaling*, *G-Protein Coupled Receptor Signaling*, and *cAMP-mediated Signaling*) (**Fig. S13a**). Within the caudate nucleus (CN), four upregulated miRNAs and four paired downregulated mRNAs could regulate four pathways (*Apoptosis Signaling*, *Axonal Guidance Signaling*, *CREB Signaling in Neurons*, and *STAT3 Signaling*) (**Fig. S13b**). Within the cerebellum (CRB), three downregulated miRNAs and two pared upregulated miRNAs could regulate two pathways (*Gap Junction Signaling* and *Neuroinflammatory Signaling*) (**Fig. S13c**).

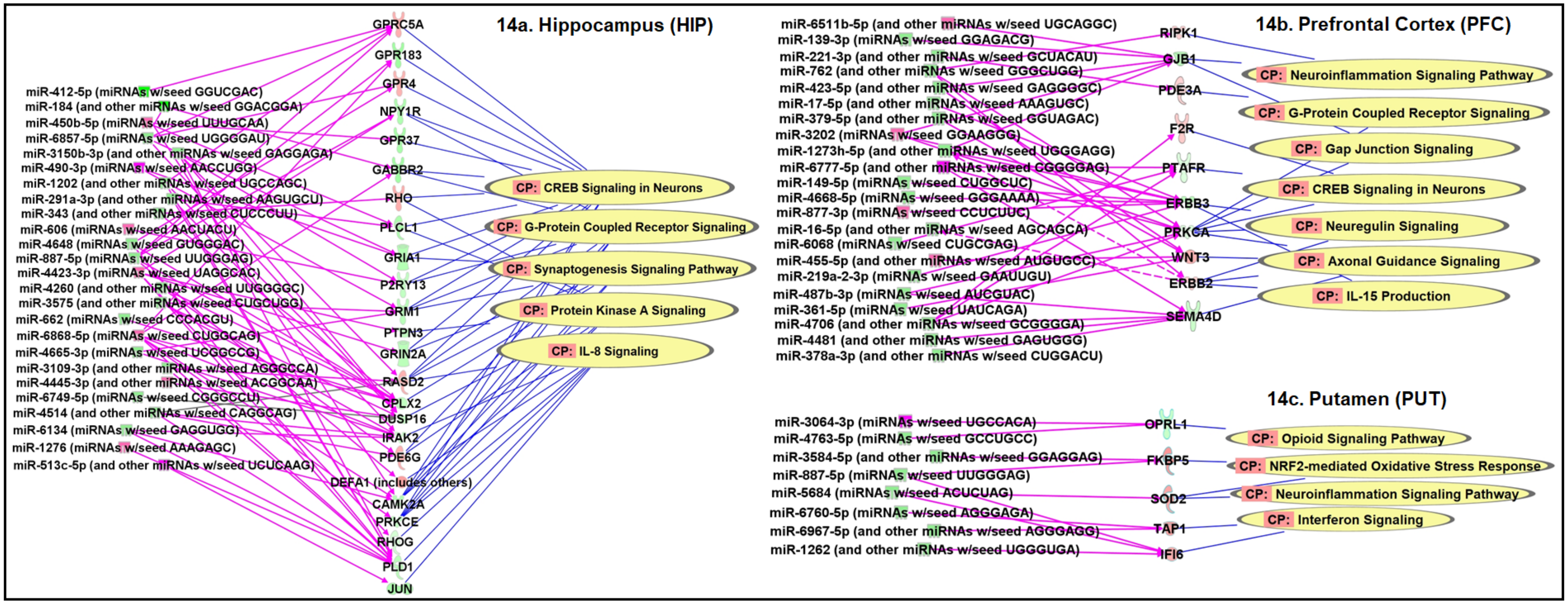


**Fig. S14** AUD-associated miRNA-mRNA regulatory networks in the hippocampus (HIPPO), the prefrontal cortex (PFC), and the putamen (PUT) of subjects with alcohol use disorder (AUD) (the Set 2 sample).

CP: Canonical pathways potentially regulated by differentially expressed [absolute fold-change (FC) > 1.3 & *P* < 0.05] and negatively correlated miRNA-mRNA pairs identified in each brain region were defined using the Ingenuity Pathway Analysis (IPA) miRNA Target Filter function. Within the hippocampus (HIP), 25 miRNAs (8 upregulated and 17 downregulated) and 24 paired mRNAs (6 upregulated and 18 downregulated) could regulate five pathways (*CREB Signaling*, *G-Protein Coupled Receptor Signaling*, *Synaptogenesis Signaling*, *Protein Kinase A Signaling*, and *IL-8 Signaling*) (**Fig. S14a**). Within the prefrontal cortex (PFC), 22 miRNAs (5 upregulated and 17 downregulated) and 10 paired mRNAs (5 upregulated and 5 downregulated) could regulate seven pathways (*Neuroinflammation Signaling*, *G-Protein Coupled Receptor Signaling*, *Gap Junction Signaling*, *CREB Signaling in Neurons*, *Neuregulin Signaling*, *Axonal Guidance Signaling*, and *IL-15 Production*) (**Fig. S14b**). Within the putamen (PUT), eight miRNAs (1 upregulated and 7 downregulated) and five paired mRNAs (4 upregulated and 1 downregulated) could regulate four pathways (*Opioid Signaling*, *NRF2-mediated Oxidative Stress Response*, *Neuroinflammation Signaling*, and *Interferon Signaling*) (**Fig. S14c**).

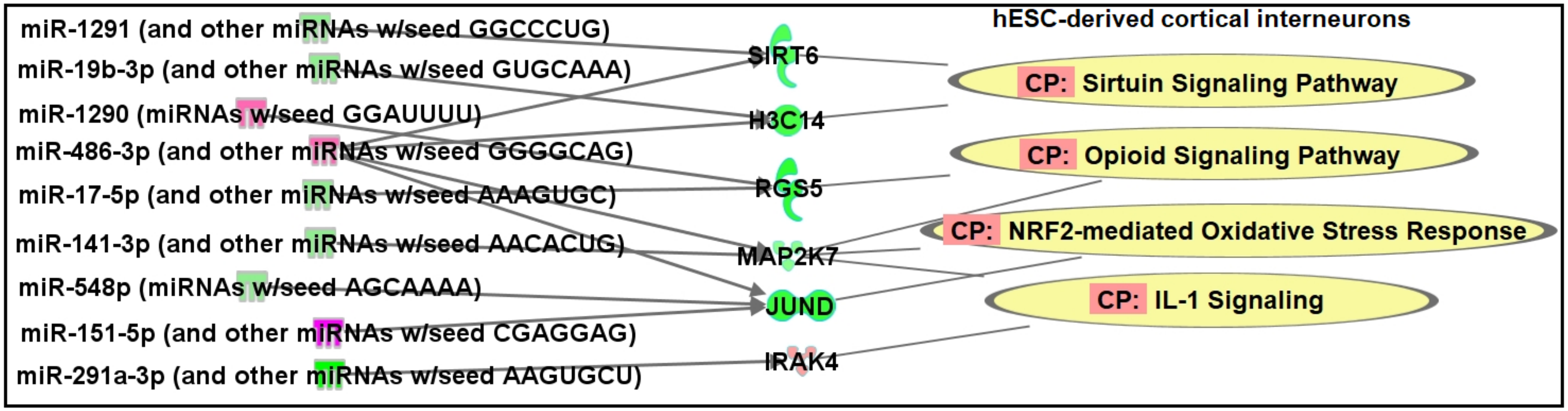


**Fig. S15** miRNA-mRNA regulatory networks in ethanol-exposed human embryonic stem cell (hESC)-derived cortical interneurons. CP: Canonical pathways potentially regulated by differentially expressed [absolute fold-change (FC) > 1.3 & $P < 0.05$] and negatively correlated miRNA-mRNA pairs were defined using the Ingenuity Pathway Analysis (IPA) miRNA Target Filter function.

**Table S1** Demographic details of brain tissue samples.

| | Set 1 | | | Set 2 | | |
|---|---|---|---|---|---|---|
| | AUD Cases | Controls | *P*-value | AUD Cases | Controls | *P*-value |
| Daily Alcohol Use in grams (Mean±SD) | 336.9 (±405.6) | 7.8 (±10.2) | 0.010 | 195 (±104.2) | 9.1 (±8.0) | 0.0002 |
| Sex | 6 (M), 6 (F) | 6 (M), 6 (F) | 1.000 | 8 (M), 0 (F) | 8 (M), 0 (F) | 1.000 |
| Age (Mean±SD) | 49 (±7) | 48 (±8) | 0.911 | 54 (±9) | 55 (±8) | 0.727 |
| Postmortem interval in hours (Mean±SD) | 37.1 (±9.4) | 29.7 (±12.6) | 0.116 | 38.6 (±19.5) | 37.1 (±14.9) | 0.865 |
| RNA interity number (Mean±SD) | AMY: 5.8 (±1.2) | AMY: 6.1 (±1.1) | 0.451 | AMY: 5.5 (±1.3) | AMY: 5.8 (±1.2) | 0.616 |
| | CN: 7.3 (±0.8) | CN: 6.8 (±1.9) | 0.453 | CN: 4.8 (±1.5) | CN: 4.9 (±1.7) | 0.881 |
| | CRB: 7.2 (±1.1) | CRB: 7.3 (±1.1) | 0.824 | CRB: 6.6 (±1.3) | CRB: 7.2 (±0.5) | 0.222 |
| | HIP: 6.2 (±1.0) | HIP: 6.0 (±1.6) | 0.790 | HIP: 6.4 (1.3) | HIP: 7.1 (0.5) | 0.167 |
| | NAc: 7.3 (±1.0) | NAc: 6.5 (±1.4) | 0.143 | NAc: N/A | NAc: N/A | N/A |
| | PFC: 6.8 (±1.0) | PFC: 6.8 (±1.6) | 0.940 | PFC: 5.3 (±1.2) | PFC: 4.9 (±1.3) | 0.467 |
| | PUT: 6.8 (±1.0) | PUT: 7.0 (±1.5) | 0.669 | PUT: 6.2 （±1.2） | PUT: 6.5 （±0.7） | 0.524 |
| | VTA: 6.1 (±0.6) | VTA: 5.9 (±1.1) | 0.603 | VTA: N/A | VTA: N/A | N/A |
| Brain weight in grams (mean±SD) | 1348.1 (±190.4) | 1416.7 (±164.9) | 0.356 | 1453.4 (±94.0) | 1505.5 (±77.4) | 0.246 |
| Brain pH (mean±SD) | 6.6 (±0.2) | 6.6 (±0.3) | 0.826 | 6.6 (±0.2) | 6.7 (±0.2) | 0.27 |
| Cerebral hemispheres | Left: 5; Right: 7 | Left: 9; Right: 3 | 0.214 | Left: 3; Right: 5 | Left: 5; Right: 3 | 0.619 |
| Smoking | Current: 10 | Current: 5 | 0.089 | Current: 5 | Current: 3 | 0.619 |
| | Never or former: 2 | Never or former: 7 | | Never or former: 3 | Never or former: 5 | |
| Liver disease | Yes: 9 | Yes: 6 | 0.400 | Yes: 6 | Yes: 7 | 1.000 |
| | No: 3 | No: 6 | | No: 2 | No: 1 | |

AUD: alcohol use disorder.

**P* values were calculated by t-tests for continuous variables and Fisher's exact tests for categorical variables.

AMY: Amygdala; CN: Caudate Nucleus; CRB: Cerebellum; HIP: Hippocampus; NAc: Nucleus Accumbens; PFC: Prefrontal Cortex; PUT: Putamen; and VTA: Ventral Tegmental Area.

Liver disease: with (Yes) or without (No) Steatosis or Congestion.

**Table S2** Small RNA-seq analysis identified miRNAs with differential expression in eight brain regions of AUD subjects (Set 1).

| Brain regions | miRNA | $log_2FC$ | AveExpr | t | *P*.Value | adj.*P*.Val | B |
|---|---|---|---|---|---|---|---|
| Amygdala (AMY) | hsa-miR-122-5p | -1.39 | 3.10 | -2.90 | 0.004 | 0.993 | -4.13 |
| | hsa-miR-6868-3p | -1.92 | 0.96 | -2.39 | 0.018 | 0.993 | -4.36 |
| | hsa-miR-412-5p | -1.04 | 4.11 | -2.10 | 0.037 | 0.993 | -4.36 |
| | hsa-miR-4326 | -1.24 | 2.93 | -2.08 | 0.039 | 0.993 | -4.39 |
| Caudate Nucleus (CN) | hsa-miR-302a-5p | -1.68 | 1.26 | -2.99 | 0.003 | 0.626 | -3.61 |
| | hsa-miR-6868-3p | -2.12 | 0.96 | -2.64 | 0.009 | 0.810 | -3.87 |
| | hsa-miR-412-5p | -1.28 | 4.11 | -2.58 | 0.011 | 0.810 | -3.73 |
| | hsa-miR-122-5p | -1.21 | 3.10 | -2.53 | 0.012 | 0.810 | -3.82 |
| | hsa-miR-144-3p | 1.20 | 3.34 | 2.26 | 0.025 | 0.810 | -4.00 |
| Cerebellum (CRB) | hsa-miR-122-5p | -1.53 | 3.10 | -3.19 | 0.002 | 0.962 | -4.03 |
| | hsa-miR-6868-3p | -1.93 | 0.96 | -2.41 | 0.017 | 0.993 | -4.35 |
| | hsa-miR-412-5p | -1.00 | 4.11 | -1.99 | 0.048 | 0.993 | -4.39 |
| Hippocampus (HIPPO) | hsa-miR-522-3p | 1.26 | 1.64 | 3.27 | 0.001 | 0.317 | -1.40 |
| | hsa-miR-10a-5p | 1.02 | 5.70 | 2.40 | 0.018 | 0.562 | -3.13 |
| | hsa-miR-1246 | 1.01 | 5.83 | 1.98 | 0.049 | 0.707 | -3.85 |
| Nucleus Accumbens (NAc) | hsa-miR-216b-5p | 1.14 | 2.14 | 2.61 | 0.010 | 1.000 | -4.25 |
| | hsa-miR-10a-5p | 1.09 | 5.70 | 2.60 | 0.010 | 1.000 | -4.15 |
| Prefrontal Cortex (PFC) | hsa-miR-486-5p | 1.00 | 8.90 | 3.27 | 0.001 | 0.519 | -2.50 |
| | hsa-miR-182-5p | 1.05 | 5.15 | 3.00 | 0.003 | 0.519 | -3.09 |
| | hsa-miR-144-5p | 1.26 | 4.27 | 2.94 | 0.004 | 0.519 | -3.22 |
| | hsa-miR-486-3p | 1.12 | 1.37 | 2.91 | 0.004 | 0.519 | -3.51 |
| | hsa-miR-5196-3p | -1.40 | 1.06 | -2.79 | 0.006 | 0.519 | -3.63 |
| | hsa-miR-4485-3p | 1.39 | 2.67 | 2.75 | 0.007 | 0.519 | -3.55 |
| | hsa-miR-144-3p | 1.28 | 3.34 | 2.40 | 0.017 | 0.675 | -3.80 |
| | hsa-miR-6868-3p | -1.69 | 0.96 | -2.11 | 0.037 | 0.815 | -4.12 |

**Table S2** (Continued)

| Brain regions | miRNA | $log_2FC$ | AveExpr | t | *P*.Value | adj.*P*.Val | B |
|---|---|---|---|---|---|---|---|
| Putamen (PUT) | hsa-miR-6868-3p | -2.63 | 0.96 | -3.29 | 0.001 | 0.360 | -1.57 |
| | hsa-miR-412-5p | -1.57 | 4.11 | -3.16 | 0.002 | 0.360 | -1.63 |
| | hsa-miR-1248 | 1.02 | 5.71 | 2.82 | 0.005 | 0.764 | -2.30 |
| | hsa-miR-5100 | 1.43 | 2.23 | 2.59 | 0.010 | 0.999 | -2.85 |
| | hsa-miR-1231 | 1.12 | 1.51 | 2.40 | 0.018 | 0.999 | -3.22 |
| Ventral tegmental Area (VTA) | hsa-miR-122-5p | -1.94 | 3.10 | -4.04 | 0.000 | 0.047 | -0.43 |
| | hsa-miR-412-5p | -1.13 | 4.11 | -2.27 | 0.024 | 1.000 | -3.50 |

Differentially expressed miRNAs with absolute $log_2FC > 1.0$ (or absolute FC > 2.0) & $P < 0.05$ are listed in the table.
$log_2FC$: the $log_2$ fold-change (FC) between cases and controls.
AveExpr: average miRNA expression level.
t: the t-statistic used to assess differential expression.
*P*.Value: the *P*-value for differential expression (this value is not adjusted for multiple testing).
adj.*P*.Val: the *P*-value adjusted for multiple testing (the default is Benjamini-Horchberg).
B: the empirical Bayes log odds of differential expression.

**Table S3** Microarray analysis identified miRNAs with differential expression in six brain regions of AUD subjects (Set 2).

| Brain regions | miRNA | $log_2$FC | AveExpr | t | *P*.Value | adj.*P*.Val | B |
|---|---|---|---|---|---|---|---|
| Amygdala (AMY) | hsa-miR-412-5p | -2.25 | 3.31 | -3.16 | 0.002 | 0.999 | -3.40 |
| Caudate Nucleus (CN) | hsa-miR-412-5p | -1.43 | 3.31 | -2.00 | 0.048 | 1.000 | -4.19 |
| Cerebellum (CRB) | hsa-miR-873-3p | -2.02 | 4.11 | -3.35 | 0.001 | 1.000 | -3.25 |
| | hsa-miR-873-5p | -1.80 | 3.38 | -3.11 | 0.003 | 1.000 | -3.44 |
| | hsa-miR-412-5p | -2.01 | 3.31 | -2.82 | 0.006 | 1.000 | -3.66 |
| Hippocampus (HIPPO) | hsa-miR-412-5p | -2.14 | 3.31 | -3.00 | 0.004 | 1.000 | -3.52 |
| | hsa-miR-184 | -2.06 | 6.85 | -2.84 | 0.006 | 1.000 | -3.65 |
| | hsa-miR-490-3p | 1.05 | 3.59 | 2.13 | 0.036 | 1.000 | -4.11 |
| | hsa-miR-1202 | -1.25 | 7.27 | -2.04 | 0.044 | 1.000 | -4.17 |
| Prefrontal Cortex (PFC) | hsa-miR-6777-5p | 1.85 | 4.90 | 6.39 | 0.000 | 0.000 | 9.60 |
| | hsa-miR-6511b-5p | 1.45 | 5.29 | 5.69 | 0.000 | 0.001 | 6.87 |
| | hsa-miR-877-3p | 1.10 | 1.73 | 4.56 | 0.000 | 0.009 | 2.75 |
| | hsa-miR-3135b | -2.87 | 10.07 | -4.39 | 0.000 | 0.012 | 2.20 |
| | hsa-miR-3202 | 1.06 | 1.68 | 3.96 | 0.000 | 0.023 | 0.79 |
| | hsa-miR-4758-5p | -1.78 | 8.33 | -3.88 | 0.000 | 0.024 | 0.54 |
| | hsa-miR-455-5p | 1.20 | 3.14 | 3.67 | 0.000 | 0.033 | -0.10 |
| | hsa-miR-4689 | 1.82 | 8.45 | 3.63 | 0.000 | 0.035 | -0.21 |
| | hsa-miR-5588-3p | -1.06 | 1.73 | -3.62 | 0.001 | 0.035 | -0.23 |
| | hsa-miR-514b-5p | 1.15 | 3.04 | 3.43 | 0.001 | 0.046 | -0.79 |
| | hsa-miR-5001-5p | -1.46 | 10.08 | -3.15 | 0.002 | 0.073 | -1.55 |
| | hsa-miR-6749-5p | -1.02 | 9.42 | -3.05 | 0.003 | 0.088 | -1.81 |
| | hsa-miR-629-3p | 1.06 | 2.99 | 2.98 | 0.004 | 0.095 | -1.98 |
| | hsa-miR-4459 | -1.16 | 8.76 | -2.92 | 0.005 | 0.105 | -2.16 |
| | hsa-miR-4668-5p | -1.72 | 8.84 | -2.86 | 0.005 | 0.115 | -2.30 |
| | hsa-miR-6068 | -1.89 | 7.94 | -2.80 | 0.006 | 0.126 | -2.45 |
| | hsa-miR-432-5p | -1.65 | 11.08 | -2.75 | 0.007 | 0.136 | -2.56 |
| | hsa-miR-423-5p | -1.81 | 9.10 | -2.73 | 0.008 | 0.139 | -2.61 |
| | hsa-miR-642a-3p | -1.23 | 4.78 | -2.72 | 0.008 | 0.140 | -2.64 |
| | hsa-miR-5584-5p | -1.42 | 3.33 | -2.71 | 0.008 | 0.140 | -2.65 |

**Table S3** (Continued)

| Brain regions | miRNA | $log_2FC$ | AveExpr | t | *P*.Value | adj.*P*.Val | B |
|---|---|---|---|---|---|---|---|
| Prefrontal Cortex (PFC) | hsa-miR-4467 | -1.49 | 9.34 | -2.71 | 0.008 | 0.140 | -2.65 |
| | hsa-miR-4745-5p | -1.66 | 9.43 | -2.66 | 0.009 | 0.151 | -2.77 |
| | hsa-miR-6779-5p | -1.71 | 8.27 | -2.66 | 0.009 | 0.151 | -2.77 |
| | hsa-miR-181b-5p | -1.31 | 12.50 | -2.61 | 0.011 | 0.160 | -2.90 |
| | hsa-miR-6800-3p | -1.14 | 3.68 | -2.57 | 0.012 | 0.166 | -2.97 |
| | hsa-miR-6787-5p | -1.32 | 7.56 | -2.53 | 0.013 | 0.175 | -3.06 |
| | hsa-miR-378h | -1.11 | 3.89 | -2.42 | 0.018 | 0.202 | -3.31 |
| | hsa-miR-338-3p | -1.27 | 4.66 | -2.41 | 0.018 | 0.205 | -3.33 |
| | hsa-miR-4461 | 1.08 | 4.69 | 2.38 | 0.019 | 0.211 | -3.38 |
| | hsa-miR-92a-3p | -1.38 | 12.08 | -2.38 | 0.020 | 0.214 | -3.40 |
| | hsa-miR-379-5p | -1.54 | 10.51 | -2.36 | 0.020 | 0.219 | -3.43 |
| | hsa-miR-361-5p | -1.04 | 11.99 | -2.35 | 0.021 | 0.221 | -3.47 |
| | hsa-miR-769-5p | -1.60 | 8.79 | -2.34 | 0.021 | 0.221 | -3.47 |
| | hsa-miR-4749-5p | -1.05 | 6.65 | -2.32 | 0.023 | 0.227 | -3.52 |
| | hsa-miR-221-3p | -1.05 | 12.01 | -2.31 | 0.023 | 0.229 | -3.54 |
| | hsa-miR-17-5p | -1.67 | 10.55 | -2.24 | 0.028 | 0.246 | -3.68 |
| | hsa-miR-149-5p | -1.12 | 11.68 | -2.21 | 0.030 | 0.253 | -3.74 |
| | hsa-miR-15b-5p | -1.28 | 8.27 | -2.19 | 0.031 | 0.258 | -3.78 |
| | hsa-miR-3613-3p | -1.25 | 8.82 | -2.19 | 0.032 | 0.260 | -3.79 |
| | hsa-miR-219a-2-3p | -1.68 | 11.26 | -2.17 | 0.033 | 0.262 | -3.82 |
| | hsa-miR-139-3p | -1.48 | 9.08 | -2.15 | 0.034 | 0.268 | -3.85 |
| | hsa-miR-487b-3p | -1.46 | 11.93 | -2.14 | 0.036 | 0.273 | -3.88 |
| | hsa-miR-421 | -1.29 | 8.68 | -2.06 | 0.042 | 0.299 | -4.02 |
| | hsa-miR-106a-5p | -1.49 | 10.37 | -2.02 | 0.046 | 0.313 | -4.10 |
| Putamen (PUT) | hsa-miR-3064-3p | 1.04 | 2.97 | 3.32 | 0.001 | 0.990 | -3.27 |
| | hsa-miR-412-5p | -1.85 | 3.31 | -2.60 | 0.011 | 1.000 | -3.82 |
| | hsa-miR-1229-5p | -1.11 | 7.80 | -2.42 | 0.018 | 1.000 | -3.94 |
| | hsa-miR-1202 | -1.32 | 7.27 | -2.16 | 0.034 | 1.000 | -4.10 |

miRNAs with absolute $log_2FC > 1.0$ (or absolute FC > 2.0) & $P < 0.05$ are listed in the table. logFC, AveExpr, t, *P*.Value, Adj.*P*.Val, and B: same as in **Table S2**.

**Table S4** Small RNA-seq analysis identified miRNAs with differential expression in hESC-derived cortical interneurons with ethanol exposure.

| miRNAs | $Log_2FC$ | AveExpr | t | *P*.Value | adj.*P*.Val | B |
|---|---|---|---|---|---|---|
| hsa-miR-151b | 1.54 | 5.43 | 2.65 | 0.039 | 0.899 | -4.44 |
| hsa-miR-151a-5p | 1.36 | 8.43 | 2.75 | 0.034 | 0.899 | -4.43 |
| hsa-miR-3135a | 1.09 | 0.17 | 4.26 | 0.006 | 0.899 | -4.35 |
| hsa-miR-548bc | -1.03 | 0.67 | -3.78 | 0.010 | 0.899 | -4.37 |
| hsa-miR-3609 | -1.18 | 0.25 | -3.66 | 0.011 | 0.899 | -4.38 |
| hsa-miR-493-5p | -1.39 | 1.90 | -3.19 | 0.019 | 0.899 | -4.40 |

Differentially expressed miRNAs with absolute $log_2FC > 1.0$ (or absolute $FC > 2.0$) & $P < 0.05$ are listed in the table.
$log_2FC$: the $log_2$ fold-change (FC) between cases and controls.
AveExpr: average miRNA expression level.
t: the t-statistic used to assess differential expression.
*P*.Value: the *P*-value for differential expression (this value is not adjusted for multiple testing).
adj.*P*.Val: the *P*-value adjusted for multiple testing (the default is Benjamini-Horchberg).
B: the empirical Bayes log odds of differential expression.

**Table S5** rRNA depletion RNA-seq analysis identified mRNAs with differential expression in eight brain regions of AUD subjects (Set 1).

| Brain regions | mRNAs | $log_2FC$ | AveExpr | t | *P*.Value | adj.*P*.Val | B |
|---|---|---|---|---|---|---|---|
| Amygdala (AMY) | *LPIN3* | 1.35 | 1.95 | 4.64 | $6.8\times10^{-6}$ | 0.036 | -1.96 |
| | *CHI3L1* | 1.72 | 3.91 | 3.82 | $1.8\times10^{-4}$ | 0.579 | -1.70 |
| | *GADD45A* | 1.03 | 3.25 | 3.61 | $3.9\times10^{-4}$ | 0.694 | -2.37 |
| | *EVI2B* | -1.37 | 0.89 | -3.57 | $4.6\times10^{-4}$ | 0.739 | -3.49 |
| | *PCDHA9* | 1.28 | 2.59 | 3.44 | $7.3\times10^{-4}$ | 0.896 | -2.97 |
| Caudate Nucleus (CN) | *FSIP2* | 1.49 | 1.21 | 4.74 | $4.4\times10^{-6}$ | 0.070 | -3.02 |
| | *EDN3* | -1.18 | 1.75 | -3.74 | $2.4\times10^{-4}$ | 0.867 | -3.42 |
| | *CYYR1* | -1.20 | 1.65 | -3.67 | $3.2\times10^{-4}$ | 0.867 | -3.55 |
| | *ABCG2* | -1.13 | 4.86 | -3.57 | $4.7\times10^{-4}$ | 0.930 | -2.44 |
| Cerebellum (CRB) | *SLIT1* | 3.10 | 6.14 | 7.95 | $2.1\times10^{-13}$ | $3.3\times10^{-9}$ | 18.27 |
| | *EPCAM* | 1.76 | 1.53 | 5.00 | $1.4\times10^{-6}$ | 0.008 | 3.78 |
| | *MAFB* | 1.46 | 2.48 | 4.97 | $1.6\times10^{-6}$ | 0.008 | 4.10 |
| | *CYTOR* | 1.38 | 1.45 | 4.77 | $3.8\times10^{-6}$ | 0.015 | 2.92 |
| | *MAGED4* | 1.09 | 4.46 | 4.53 | $1.1\times10^{-5}$ | 0.030 | 3.01 |
| | *CCDC158* | 1.32 | 0.93 | 4.27 | $3.2\times10^{-5}$ | 0.056 | 1.12 |
| | *COLCA2* | 1.26 | 0.98 | 4.04 | $7.8\times10^{-5}$ | 0.104 | 0.48 |
| | *TMEM176A* | 1.11 | 2.33 | 3.90 | $1.4\times10^{-4}$ | 0.130 | 0.58 |
| | *DACH2* | 1.47 | 2.92 | 3.84 | $1.7\times10^{-4}$ | 0.137 | 0.57 |
| | *SLC2A4* | 1.47 | 0.83 | 3.82 | $1.9\times10^{-4}$ | 0.137 | -0.15 |
| | *AZGP1* | 1.42 | 1.34 | 3.73 | $2.6\times10^{-4}$ | 0.179 | -0.19 |
| | *PCDH8* | 1.73 | 4.45 | 3.64 | $3.6\times10^{-4}$ | 0.237 | 0.01 |
| | *CYYR1* | -1.21 | 1.65 | -3.63 | $3.7\times10^{-4}$ | 0.238 | -0.46 |
| | *RAMP2* | -1.15 | 1.90 | -3.56 | $4.7\times10^{-4}$ | 0.271 | -0.55 |
| | *ALDH7A1P1* | 1.03 | 1.23 | 3.52 | $5.4\times10^{-4}$ | 0.271 | -0.77 |

**Table S5** (Continued)

| Brain regions | mRNAs | $\log_2$FC | AveExpr | t | *P*.Value | adj.*P*.Val | B |
|---|---|---|---|---|---|---|---|
| Cerebellum (CRB) | *ABHD12B* | 1.22 | 1.48 | 3.52 | $5.4\times10^{-4}$ | 0.271 | -0.53 |
| | *B4GALNT3* | 1.06 | 2.85 | 3.50 | $6.0\times10^{-4}$ | 0.277 | -0.48 |
| | *PLPP4* | -1.30 | 2.60 | -3.49 | $6.1\times10^{-4}$ | 0.277 | -0.55 |
| | *MT1G* | 1.55 | 1.36 | 3.47 | $6.5\times10^{-4}$ | 0.280 | -0.78 |
| | *DCAF12L2* | -1.24 | 0.67 | -3.46 | $6.8\times10^{-4}$ | 0.285 | -1.04 |
| | *SPSB4* | 1.02 | 0.72 | 3.40 | $8.3\times10^{-4}$ | 0.321 | -1.18 |
| Hippocampus (HIPPO) | *CDH7* | 1.50 | 3.39 | 4.44 | $1.6\times10^{-5}$ | 0.095 | 1.37 |
| | *CCNO* | 1.93 | 0.91 | 4.41 | $1.8\times10^{-5}$ | 0.095 | -0.17 |
| | *FAM83G* | 1.26 | 0.49 | 3.90 | $1.4\times10^{-4}$ | 0.332 | -1.64 |
| | *TNK1* | 1.37 | 1.37 | 3.80 | $2.0\times10^{-4}$ | 0.332 | -0.98 |
| | *CYS1* | 1.01 | 2.15 | 3.75 | $2.4\times10^{-4}$ | 0.332 | -1.07 |
| | *RGS16* | 1.41 | 2.13 | 3.53 | $5.3\times10^{-4}$ | 0.541 | -1.48 |
| | *GRM8* | 1.02 | 3.46 | 3.49 | $6.0\times10^{-4}$ | 0.541 | -0.98 |
| | *DMKN* | 1.26 | 2.46 | 3.46 | $6.8\times10^{-4}$ | 0.541 | -1.31 |
| | *TMEM200B* | 1.29 | 1.22 | 3.43 | $7.5\times10^{-4}$ | 0.541 | -1.79 |
| | *CYYR1* | -1.11 | 1.65 | -3.39 | $8.5\times10^{-4}$ | 0.541 | -1.99 |
| | *EDN3* | -1.06 | 1.75 | -3.39 | $8.5\times10^{-4}$ | 0.541 | -1.87 |
| Nucleus Accumbens (NAc) | *FSIP2* | 1.32 | 1.21 | 4.19 | $4.4\times10^{-5}$ | 0.512 | -3.45 |
| | *EDN3* | -1.19 | 1.75 | -3.72 | $2.7\times10^{-4}$ | 1.000 | -3.54 |
| | *TMEM255A* | 1.00 | 3.05 | 3.72 | $2.7\times10^{-4}$ | 1.000 | -2.88 |
| | *CA4* | -1.08 | 2.41 | -3.55 | $5.0\times10^{-4}$ | 1.000 | -3.30 |
| Prefrontal Cortex (PFC) | *ACP7* | -1.60 | 1.06 | -4.72 | $4.9\times10^{-6}$ | 0.054 | 2.85 |
| | *GJB1* | -2.06 | 3.19 | -4.64 | $6.7\times10^{-6}$ | 0.054 | 3.25 |
| | *KLK6* | -2.07 | 3.70 | -4.20 | $4.2\times10^{-5}$ | 0.124 | 1.78 |
| | *UGT8* | -1.70 | 5.75 | -4.17 | $4.7\times10^{-5}$ | 0.124 | 1.78 |

**Table S5** (Continued)

| Brain regions | mRNAs | $log_2FC$ | AveExpr | t | *P*.Value | adj.*P*.Val | B |
|---|---|---|---|---|---|---|---|
| Prefrontal Cortex | *CA14* | -1.66 | 1.91 | -4.13 | $5.5\times10^{-5}$ | 0.124 | 1.16 |
| (PFC) | *ST18* | -1.49 | 6.52 | -4.13 | $5.6\times10^{-5}$ | 0.124 | 1.63 |
| | *MYOT* | -1.64 | 0.80 | -4.02 | $8.7\times10^{-5}$ | 0.124 | 0.33 |
| | *RNASE1* | -1.35 | 5.15 | -4.01 | $8.9\times10^{-5}$ | 0.124 | 1.22 |
| | *SMIM5* | -1.86 | 1.67 | -3.99 | $1.0\times10^{-4}$ | 0.124 | 0.70 |
| | *TGFBI* | 1.77 | 1.86 | 3.96 | $1.1\times10^{-4}$ | 0.124 | 0.63 |
| | *EVI2A* | -1.77 | 4.11 | -3.96 | $1.1\times10^{-4}$ | 0.124 | 1.02 |
| | *CLCA4* | -1.55 | 1.49 | -3.92 | $1.3\times10^{-4}$ | 0.128 | 0.43 |
| | *FOLH1* | -1.61 | 4.00 | -3.90 | $1.4\times10^{-4}$ | 0.129 | 0.83 |
| | *ANLN* | -1.66 | 5.79 | -3.87 | $1.6\times10^{-4}$ | 0.132 | 0.75 |
| | *NIPAL4* | -1.61 | 1.63 | -3.85 | $1.6\times10^{-4}$ | 0.132 | 0.35 |
| | *CNDP1* | -1.60 | 6.14 | -3.85 | $1.7\times10^{-4}$ | 0.132 | 0.69 |
| | *PLLP* | -1.24 | 5.03 | -3.79 | $2.0\times10^{-4}$ | 0.156 | 0.51 |
| | *CYB5R2* | -1.30 | 2.39 | -3.76 | $2.3\times10^{-4}$ | 0.162 | 0.21 |
| | *TTYH2* | -1.14 | 5.97 | -3.76 | $2.3\times10^{-4}$ | 0.162 | 0.40 |
| | *PRIMA1* | -1.38 | 3.81 | -3.67 | $3.2\times10^{-4}$ | 0.175 | 0.09 |
| | *FA2H* | -1.60 | 4.30 | -3.66 | $3.3\times10^{-4}$ | 0.175 | 0.09 |
| | *ICOSLG* | -1.34 | 4.68 | -3.63 | $3.7\times10^{-4}$ | 0.181 | 0.00 |
| | *COL4A5* | -1.24 | 5.32 | -3.62 | $3.8\times10^{-4}$ | 0.181 | -0.03 |
| | *MYRF* | -1.51 | 6.86 | -3.62 | $3.8\times10^{-4}$ | 0.181 | -0.03 |
| | *SLC45A3* | -1.54 | 2.97 | -3.62 | $3.9\times10^{-4}$ | 0.181 | -0.11 |
| | *RASGRP3* | -1.34 | 4.18 | -3.60 | $4.1\times10^{-4}$ | 0.181 | -0.09 |
| | *PLPP2* | -1.30 | 2.63 | -3.58 | $4.5\times10^{-4}$ | 0.188 | -0.27 |
| | *KCNH8* | -1.20 | 4.51 | -3.58 | $4.5\times10^{-4}$ | 0.188 | -0.17 |
| | *TRIM59* | -1.60 | 2.83 | -3.54 | $5.2\times10^{-4}$ | 0.196 | -0.37 |

**Table S5** (Continued)

| Brain regions | mRNAs | $log_2FC$ | AveExpr | t | *P*.Value | adj.*P*.Val | B |
|---|---|---|---|---|---|---|---|
| Prefrontal Cortex | *IP6K3* | -1.83 | 1.11 | -3.54 | $5.2\times10^{-4}$ | 0.196 | -0.65 |
| (PFC) | *SLC38A5* | -1.19 | 2.80 | -3.52 | $5.5\times10^{-4}$ | 0.204 | -0.47 |
| | *HSD11B1* | -1.48 | 1.16 | -3.51 | $5.7\times10^{-4}$ | 0.204 | -0.86 |
| | *PLP1* | -1.45 | 10.28 | -3.50 | $5.9\times10^{-4}$ | 0.204 | -0.41 |
| | *ERBB3* | -1.22 | 5.48 | -3.50 | $5.9\times10^{-4}$ | 0.204 | -0.40 |
| | *ASPA* | -1.27 | 5.03 | -3.49 | $6.1\times10^{-4}$ | 0.204 | -0.42 |
| | *FSIP2* | 1.11 | 1.21 | 3.49 | $6.1\times10^{-4}$ | 0.204 | -0.89 |
| | *SLC5A11* | -1.77 | 2.68 | -3.47 | $6.6\times10^{-4}$ | 0.209 | -0.57 |
| | *PRR18* | -1.18 | 4.20 | -3.45 | $6.9\times10^{-4}$ | 0.216 | -0.53 |
| | *NKX6-2* | -1.47 | 3.57 | -3.38 | $8.9\times10^{-4}$ | 0.236 | -0.75 |
| | *SGK2* | -1.20 | 2.50 | -3.38 | $9.0\times10^{-4}$ | 0.236 | -0.85 |
| | *ELOVL1* | -1.12 | 4.55 | -3.37 | $9.4\times10^{-4}$ | 0.236 | -0.79 |
| | *HHIP* | -1.36 | 5.27 | -3.36 | $9.4\times10^{-4}$ | 0.236 | -0.79 |
| | *SH3TC2* | -1.31 | 5.05 | -3.36 | $9.5\times10^{-4}$ | 0.236 | -0.80 |
| | *PRR5L* | -1.11 | 3.05 | -3.36 | $9.5\times10^{-4}$ | 0.236 | -0.85 |
| | *CTNNA3* | -1.38 | 5.50 | -3.36 | $9.6\times10^{-4}$ | 0.236 | -0.81 |
| | *MOG* | -1.46 | 6.18 | -3.35 | $9.9\times10^{-4}$ | 0.240 | -0.84 |
| Putamen | *MAFF* | 1.53 | 1.82 | 4.01 | $8.8\times10^{-5}$ | 0.270 | -0.62 |
| (PUT) | *FAM110C* | 1.46 | 1.52 | 3.98 | $1.0\times10^{-4}$ | 0.270 | -0.60 |
| | *PLCD4* | 1.00 | 2.36 | 3.91 | $1.3\times10^{-4}$ | 0.285 | -0.62 |
| | *CHI3L1* | 1.72 | 3.91 | 3.82 | $1.8\times10^{-4}$ | 0.285 | 0.02 |
| | *FSIP2* | 1.20 | 1.21 | 3.80 | $2.0\times10^{-4}$ | 0.285 | -1.47 |
| | *EDN3* | -1.19 | 1.75 | -3.78 | $2.2\times10^{-4}$ | 0.289 | -1.21 |
| | *SERPINA3* | 2.74 | 2.96 | 3.73 | $2.6\times10^{-4}$ | 0.301 | -0.47 |
| | *MT1X* | 1.57 | 3.47 | 3.62 | $3.8\times10^{-4}$ | 0.369 | -0.69 |

**Table S5** (Continued)

| Brain regions | mRNAs | $log_2FC$ | AveExpr | t | *P*.Value | adj.*P*.Val | B |
|---|---|---|---|---|---|---|---|
| Putamen (PUT) | *MDK* | 1.06 | 2.67 | 3.61 | $3.9×10^{-4}$ | 0.369 | -1.08 |
| | *CEBPD* | 1.49 | 3.14 | 3.42 | $7.9×10^{-4}$ | 0.515 | -1.23 |
| | *C1QL3* | 1.74 | 2.59 | 3.40 | $8.3×10^{-4}$ | 0.515 | -1.22 |
| Ventral Tegmental Area (VTA) | *NLRP2* | -1.92 | 1.35 | -3.91 | $1.3×10^{-4}$ | 0.414 | -2.63 |
| | *CHI3L1* | 1.69 | 3.91 | 3.76 | $2.3×10^{-4}$ | 0.517 | -1.24 |
| | *GNA14* | 1.16 | 2.45 | 3.74 | $2.5×10^{-4}$ | 0.517 | -2.04 |
| | *LOXL2* | 1.14 | 1.87 | 3.67 | $3.2×10^{-4}$ | 0.540 | -2.62 |
| | *MAFB* | 1.03 | 2.48 | 3.58 | $4.5×10^{-4}$ | 0.540 | -2.34 |
| | *NPIPB15* | 2.09 | 0.57 | 3.50 | $5.8×10^{-4}$ | 0.540 | -3.30 |

Differentially expressed coding genes (or mRNAs) with absolute $log_2FC > 1.0$ (or absolute FC > 2.0) & $P < 0.001$ are listed in the table.

$log_2FC$: the $log_2$ fold-change (FC) between cases and controls.

AveExpr: average miRNA expression level.

t: the t-statistic used to assess differential expression.

*P*.Value: the *P*-value for differential expression (this value is not adjusted for multiple testing).

adj.*P*.Val: the *P*-value adjusted for multiple testing (the default is Benjamini-Horchberg).

B: the empirical Bayes log odds of differential expression.

**Table S6** Microarray analysis identified mRNAs with differential expression in six brain regions of AUD subjects (Set 2).

| Brain regions | mRNAs | logFC | AveExpr | t | *P*.Value | adj.*P*.Val | B |
|---|---|---|---|---|---|---|---|
| Amygdala (AMY) | *OCLN* | 1.28 | 5.66 | 5.45 | $4.6\times10^{-7}$ | 0.031 | -0.71 |
| | *GLS2* | -1.09 | 5.50 | -3.65 | $4.5\times10^{-4}$ | 1.000 | -2.72 |
| Caudate Nucleus (CN) | N/A | | | | | | |
| Cerebellum (CRB) | N/A | | | | | | |
| Hippocampus (HIPPO) | *SLC47A1* | -1.17 | 5.15 | -4.69 | $1.0\times10^{-5}$ | 0.273 | -1.44 |
| | *PROX1* | -1.16 | 6.57 | -3.95 | $1.6\times10^{-4}$ | 0.711 | -2.30 |
| | *MYO5B* | -1.03 | 4.50 | -3.93 | $1.7\times10^{-4}$ | 0.724 | -2.32 |
| | *TNC* | -1.13 | 4.37 | -3.60 | $5.3\times10^{-4}$ | 0.844 | -2.68 |
| Prefrontal Cortex (PFC) | N/A | | | | | | |
| Putamen (PUT) | *OCLN* | 1.02 | 5.66 | 4.33 | $3.9\times10^{-5}$ | 1.000 | -2.00 |

Differentially expressed coding genes (or mRNAs) with absolute $\log_2$FC > 1.0 (or absolute FC > 2.0) & $P < 0.001$ are listed in the table.
$\log_2$FC: the $\log_2$ fold-change (FC) between cases and controls.
AveExpr: average miRNA expression level.
t: the t-statistic used to assess differential expression.
*P*.Value: the *P*-value for differential expression (this value is not adjusted for multiple testing).
adj.*P*.Val: the *P*-value adjusted for multiple testing (the default is Benjamini-Horchberg).
B: the empirical Bayes log odds of differential expression.

**Table S7** mRNA-seq analysis identified mRNAs with differential expression in ethanol-exposed hESC-derived cortical interneurons.

| mRNAs | logFC | AveExpr | t | *P*.Value | adj.*P*.Val | B |
|---|---|---|---|---|---|---|
| *H3C13* | -1.42 | 3.51 | -4.75 | $2.2\times10^{-3}$ | 1.000 | -4.36 |
| *NUBP2* | -1.31 | 2.74 | -4.57 | $2.7\times10^{-3}$ | 1.000 | -4.37 |
| *MIF* | -1.22 | 5.20 | -4.49 | $3.0\times10^{-3}$ | 1.000 | -4.37 |
| *FAM171A2* | -1.22 | 3.59 | -4.37 | $3.4\times10^{-3}$ | 1.000 | -4.37 |
| *DPM3* | -1.11 | 2.63 | -4.05 | $5.1\times10^{-3}$ | 1.000 | -4.39 |
| *H2AW* | -1.37 | 3.10 | -3.93 | $5.9\times10^{-3}$ | 1.000 | -4.39 |
| *ABHD17A* | -1.16 | 3.57 | -3.91 | $6.1\times10^{-3}$ | 1.000 | -4.39 |
| *ZNF219* | -1.41 | 3.11 | -3.81 | $6.9\times10^{-3}$ | 1.000 | -4.40 |
| *H3C14* | -1.16 | 5.56 | -3.79 | $7.1\times10^{-3}$ | 1.000 | -4.40 |
| *H3C15* | -1.16 | 5.56 | -3.79 | $7.1\times10^{-3}$ | 1.000 | -4.40 |
| *DLX2* | -1.11 | 4.31 | -3.77 | $7.2\times10^{-3}$ | 1.000 | -4.40 |
| *LMTK3* | -1.01 | 2.43 | -3.75 | $7.4\times10^{-3}$ | 1.000 | -4.40 |
| *ZGPAT* | -1.02 | 2.90 | -3.72 | $7.8\times10^{-3}$ | 1.000 | -4.40 |
| *SEPTIN7-DT* | -1.15 | 2.61 | -3.69 | $8.0\times10^{-3}$ | 1.000 | -4.40 |
| *PLPPR3* | -1.16 | 4.19 | -3.63 | $8.7\times10^{-3}$ | 1.000 | -4.40 |
| *XRCC3* | -1.11 | 2.27 | -3.62 | $8.9\times10^{-3}$ | 1.000 | -4.40 |
| *DLGAP3* | -1.01 | 2.14 | -3.55 | $9.7\times10^{-3}$ | 1.000 | -4.41 |
| *FGF14-IT1* | -1.05 | 2.68 | -3.53 | $9.9\times10^{-3}$ | 1.000 | -4.41 |
| *MZT2B* | -1.24 | 3.83 | -3.53 | $9.9\times10^{-3}$ | 1.000 | -4.41 |

Differentially expressed coding genes (or mRNAs) with absolute $\log_2$FC (or absolute FC > 2.) > 1.0 & $P < 0.01$ are listed in the table.
$\log_2$FC: the $\log_2$ fold-change (FC) between cases and controls.
AveExpr: average miRNA expression level.
t: the t-statistic used to assess differential expression.
*P*.Value: the *P*-value for differential expression (this value is not adjusted for multiple testing).
adj.*P*.Val: the *P*-value adjusted for multiple testing (the default is Benjamini-Horchberg).
B: the empirical Bayes log odds of differential expression.

**Table S8** Canonical pathways potentially regulated by differentially expressed and negatively correlated miRNA-mRNA pairs.

| | Set 1 (RNA-seq) | | | | | | | | Set 2 (Affymetrix array) | | | | | | hESC-derived cortical interneurons |
|---|---|---|---|---|---|---|---|---|---|---|---|---|---|---|---|
| Canonical Pathways | AMY | CN | CRB | HIP | NAc | PFC | PUT | VTA | AMY | CN | CRB | HIP | PFC | PUT | |
| CREB Signaling in Neurons | √ | √ | √ | √ | √ | √ | | √ | √ | √ | | √ | √ | | |
| IL-8 Signaling | √ | | | | | √ | √ | √ | | | | √ | | | |
| Axonal Guidance Signaling | √ | √ | | √ | | √ | | | √ | √ | | | √ | | |
| Neuroinflammation Signaling | | √ | √ | | | | | | | | √ | √ | √ | √ | |
| Synaptogensis Signaling Pathway | | | √ | √ | | | | | √ | | | √ | √ | | |
| G-Protein Coupled Receptor Signaling | | | | √ | | √ | | | √ | | | √ | √ | | |
| Gap Junction Signaling | | √ | | | | | | √ | | | √ | | √ | | |
| Opioid Signaling Pathway | | | | | | | | √ | √ | | | | | √ | √ |
| NRF-mediated Oxidative Stress Response | | | | | | | | √ | | | | | | √ | √ |
| STAT3 Pathway | √ | | | | | | | | | √ | | | | | |
| Sirtuin Signaling | | | | | | | √ | | | | | | | | √ |
| Protein Kinase A Signaling | | | | | | | | | √ | | | √ | | | |
| cAMP-mediated signaling | | | | | | | | | √ | | | | | | |
| Apoptosis Signaling | | | | | | | | | | √ | | | | | |
| Neuregulin Signaling | | | | | | | | | | | | | √ | | |
| Interferon Sigaling | | | | | | | | | | | | | | √ | |
| IL-1 Signaling | | | | | | | | | | | | | | | √ |

AMY: Amygdala; CN: Caudate Nucleus; CRB: Cerebellum; HIP: Hippocampus; NAc: Nucleus Accumbens; PFC: Prefrontal Cortex; PUT: Putamen; and VTA: Ventral Tegmental Area.

Set 1 (RNA-seq): miRNA and mRNA expression profiles in eight brain regions (Set 1 sample) were analyzed by RNA-seq.

Set 2 (Affymetrix array): miRNA and mRNA expression profiles in six brain regions (Set 2 sample) were analyzed by Affymetrix expression array assay.

hESC-derived cortical interneurons: miRNA and mRNA expression profiles in hESC-derived cortical interneurons were analyzed by RNA-seq.

"√" means that a specific canonical pathway is potentially regulated by differentially expressed and negatively correlated miRNA-mRNA pairs in a specific brain region of subjects with alcohol use disorder (AUD) or ethanol-exposed hESC-derived cortical interneurons.
